# Can a Conference be Epidemiologically Conscious? A Pilot Study of Implementing COVID-19 Mitigation Measures at an In-Person Conference

**DOI:** 10.1101/2025.06.30.25328802

**Authors:** Lorenzo Servitje, Thomas McAndrew, Kathleen Bachynski, Chrysan Cronin, Martha Lincoln, Jessecae K. Marsh, Ashlee Simon, Lily Kaganov, Brooke Erickson

## Abstract

**Introduction**: Public health guidance about respiratory agents often focuses on the county level, overlooking local events that may increase transmission—such as conferences. **Objectives:** To understand conference-level mitigations, we assessed the use of a “COVID Conscious” protocol implemented during the 2022 North American Victorian Studies Association conference (Bethlehem, PA; September 28th to October 2nd). **Methods:** Eleven surveys measured attendees’ mitigation behaviors before, during, and after the conference. The proposed conference mitigation strategy included—for all 258 attendees (84 completed surveys)—provision of N95 masks, access to rapid antigen tests (RATs) and local public health information, and air quality improvement. **Results:** The proportion of attendees testing was highest in the two days after the conference (85% and 88%) with those reporting symptoms having higher odds of testing (OR: 3.81; 95CI% [1.19, 7.63; *p<*0.01). The proportion of attendees masking was approximately 90% throughout the conference. Respondents endorsed the provision of N95 masks (92%) and RATs (89%) and rated local surveillance information as the least helpful mitigation (14%). Two cases of COVID-19 were reported within seven days of the conference ending. **Discussion:** Our findings highlight an opportunity for critical public health practice: considering the convergence of “publics” during conferences and accounting for their unique structural inequities. Conference-level mitigation may act as a public health intervention by potentially reducing the infectious disease burden conferences introduce to a community, its local population and occupational health. To this end, we present a protocol for organizers to consider for implementation and further study

## Background

A return to in-person events such as academic conferences challenges event organizers and attendees seeking to limit the risk of respiratory disease transmission ( Dupouy et al., 2023; Ehteshami et al., 2023; Gomes et al., 2022; Gottlieb et al., 2020; Silver et al., 2022; Sumer et al., 2022; Talcott et al., 2022). Past work shows transmission can be minimized by implementing masking (Huang et al., 2022; Suh et al., 2022; Talic et al., 2021), ventilation and air purification (Aldekheel et al., 2022; Di Gilio et al., 2021; Morawska et al., 2020), testing (Brook et al., 2021), and vaccination (Lipsitch & Dean, 2020; 2022; Stokel-Walker, 2022).

In the past, the Centers for Disease Control and Prevention (CDC) has recommended implementing personal and community-level COVID-19 interventions based on county-level hospital admission levels (CDC, 2022a; Christie et al., 2021; CDC, 2023). However, there has been no guidance specific to events like conferences. This study aimed to implement a conferencing protocol to mitigate the spread of SARS-CoV-2 and assess the acceptability of this protocol to attendees during the Fall of 2022 in the United States.

Conference populations are unique because of the sustained close contact of attendees and the additional risk that a conference may pose to the different local populations and venue employees who share spaces with attendees throughout the conference (Green et al., 2022). There have been numerous reports of conferences (including in biomedical science and public health) becoming superspreader events (Ehteshami et al., 2023; Gomes et al., 2022; Silver et al., 2022; Sumer et al., 2022; Talcott et al., 2021; Swaminathan et al., 2022). Notable conference outbreaks include the CDC’s Epidemic Intelligence Service (EIS) Conference, which reported 181 cases of COVID-19 among in-person attendees (∼10% of the conference population) (CDC, 2023).

To mitigate superspreader events, past research has studied the uptake of preventive measures and test positivity at conferences and found that providing attendees with multiple mitigation measures may reduce the risk of disease transmission (Dupouy et al., 2023; Ehteshami et al., 2023; Gomes et al., 2022; Silver et al., 2022; Sumer et al., 2022; Talcott et al., 2021). However, mitigation strategies differ by conference as do the demographics and perceptions of risk among attendees. (Sumer et al., 2022). Existing studies on conference mitigations and uptake have suggested that high rates of adherence to mandated protocols might be due to a selection bias in their samples (medical, scientific, and infectious disease experts) (Ehteshami et al., 2023; Sumer et al., 2022).

Despite some organizers’ efforts to document incident cases among conference attendees and implement protocols to prevent transmission at conferences, there remains little work on the uptake and impact of mitigation strategies at conferences and the communities that host them. The paucity of guidelines and research on academic conferences during the COVID-19 pandemic highlights a gap and an occasion for critical public health reflection and practice. Public health guidance for these venues was relegated to something between a ‘medicalized’ responsibility for individuals and DIY project for organizers who may lack expertise (Goldberg, 2021; Schrecker, 2022). Bell and Green (2015) suggest that one inflection of criticality vis-a-vis public health entails ‘turn[ing] the spotlight on public health practice itself’. Two elements that Schrecker (2022) posits for engaged scholarship and practice of critical public health are i) not only a description of but a commitment to ‘the idea that many health inequalities in health outcomes are ethically indefensible and ii) attention to the social arrangements and institutions that shape health inequities (p. 139-140). Following the first, we present a detailed protocol implemented at an academic conference that aimed to mitigate the potential increase in transmission the event might have local community, host venues, and conference attendees. We collected survey data on attendees’ reactions to this protocol and longitudinal data on the percentage of attendees who tested for COVID-19 and masked to prevent the pathogen’s spread. Following the second, we discuss how conferences themselves can manifest as institutions and social arrangements that affect existing social and health inequalities but, in that same capacity, also provide an opportunity for critical public health practice.

## Methods

### Timeline and Venue

The North American Victorian Studies (NAVSA) Conference was held between September 29th and October 2nd, 2022 in Bethlehem, PA (USA) in Northampton County (four days total). It was attended by a total of 258 in-person and 15 remote participants. Between the weeks of September 24th and October 8th, the bivalent mRNA vaccines had just become widely available (authorized on September 1, 2022, for ages 12 and older) (Food and Drug Administration, 2022). The CDC had changed surveillance indicators from community transmission to ‘Community Levels’ six months prior to the start of the conference. NAVSA 2022 was the organization’s first in-person conference since 2019 and was held at a local hotel for three days and at Lehigh University for one day.

### Conference Protocol

NAVSA organizers implemented and promoted mitigation methods with demonstrated effectiveness in minimizing COVID-19 transmission (Aldekheel et al., 2022; Brook et al., 2021; Di Gilio et al., 2021; Morawska et al., 2020; Talic et al., 2021). Prior to the conference, all registered attendees were provided with NAVSA’s ‘COVID Conscious’ protocol (Supplement A). The ‘COVID Conscious’ protocol was intended not only to keep attendees safe, but also to reduce the event’s ‘epidemiological footprint’— its potential impact on local respiratory infection trends—and reduce the risk of infection by and for attendees during travel and upon returning home.

Attendees were not mandated to change their behavior but encouraged to adhere to the following guidelines:

1. Masking indoors.
2. Testing: before traveling, in the morning before attending events, or when feeling ill.
3. Isolating in the event of a positive test during the conference.
4. Following standard precautions such as handwashing and distancing (NAVSA, 2022).

Registered conference attendees were notified via email on August 31st, 2022 of the following measures the organizers would implement at the conference:

1. Provide attendees with free N95 masks and rapid antigen tests (RATs) before and during the conference. Prior to the conference, attendees could request two N95 masks and one RAT via mail.
2. Provide standard guidance about prevention measures and updated community transmission rates in Northampton County.
3. Indoor Air quality (IAQ): Monitor room air exchange (ACH) rates with CO2 monitors and implement auxiliary air purification with Corsi-Rosenthal Boxes (Figure S1) if the rates fell below 5 ACH (Allen & Ibrahim, 2021). (See Supplement B for discussion of air purification; Figure S2 for example room data.)
4. Reduce attendee density with hybrid option: 20% of the panels had Zoom functionality. The hybrid option reduced in-person attendance by 15 people total, approximately 5% of the conference attendance (15/273, assuming the 15 people would have attended the conference, raising the population from 258 to 273).
5. Provide refunds and financial assistance to attendees who tested positive before or during the conference or had to cancel for medical reasons (NAVSA, 2022).

A complete list of policies and procedures is available in Supplement A.

### Participants, Survey Design, and Measurement

All 258 conference attendees were made aware of this study and 84 (33%) elected to participate. Survey questions covered uptake of RAT and mask use, mitigation use and behaviors, and test positivity before, during, and after the conference (See Figure 1). Written informed consent was provided in the first pre-conference survey. Registrants were notified that access to tests and masks was free and not contingent on study participation. Participants were sent: (i) one pre-conference survey; (ii) up to four daily check-in surveys during the conference (depending on their reported first/last attendance days), (iii) one post-conference debrief survey, and (iv) five post-conference check-in surveys. In order to provide an additional mechanism to report positive tests, a post-conference positivity survey was sent following the conference (the survey was open Nov. 29 to Dec. 11) to all conference attendees, whether or not they participated in the study. All surveys were administered via Qualtrics. (See Supplement C for survey questions). Participants were deidentified for data analysis.

**FIG. 1.**
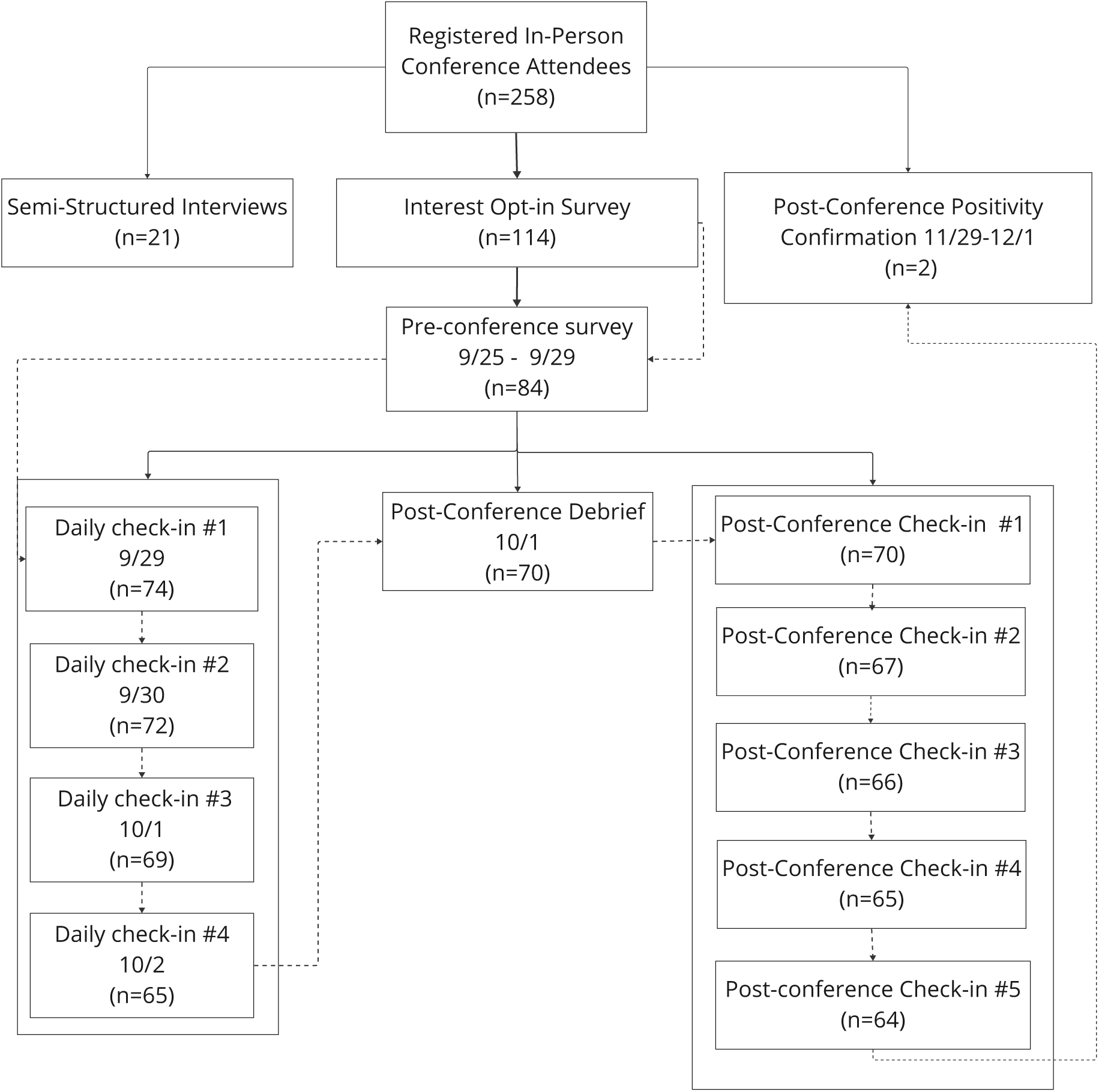
Study flow diagram. Dashed lines indicate the ideal survey completion pathway for a participant that completes every survey: interest/opt-in; pre-conference survey; attends all days of the conference and completes all daily check-ins; completes post-conference debrief; completes all post-conference check-ins. A total of 33/84 (39%) participants completed the series in this manner. Not all attendees were able to complete 4 daily check-ins during the conference, as it required staying for the entire duration of the conference (including a professionalization workshop for graduate students on the last day). A total of 62/84 (74%) completed nine or more surveys. See Figure 4 for breakdown of survey participation by number and proportion.

The pre-survey collected participants’ sociodemographic information, perceived health status, self-reported risk factors associated with severe illness, COVID-19 vaccination status, and previous COVID-19 infection (Table 1).

**TABLE 1:**
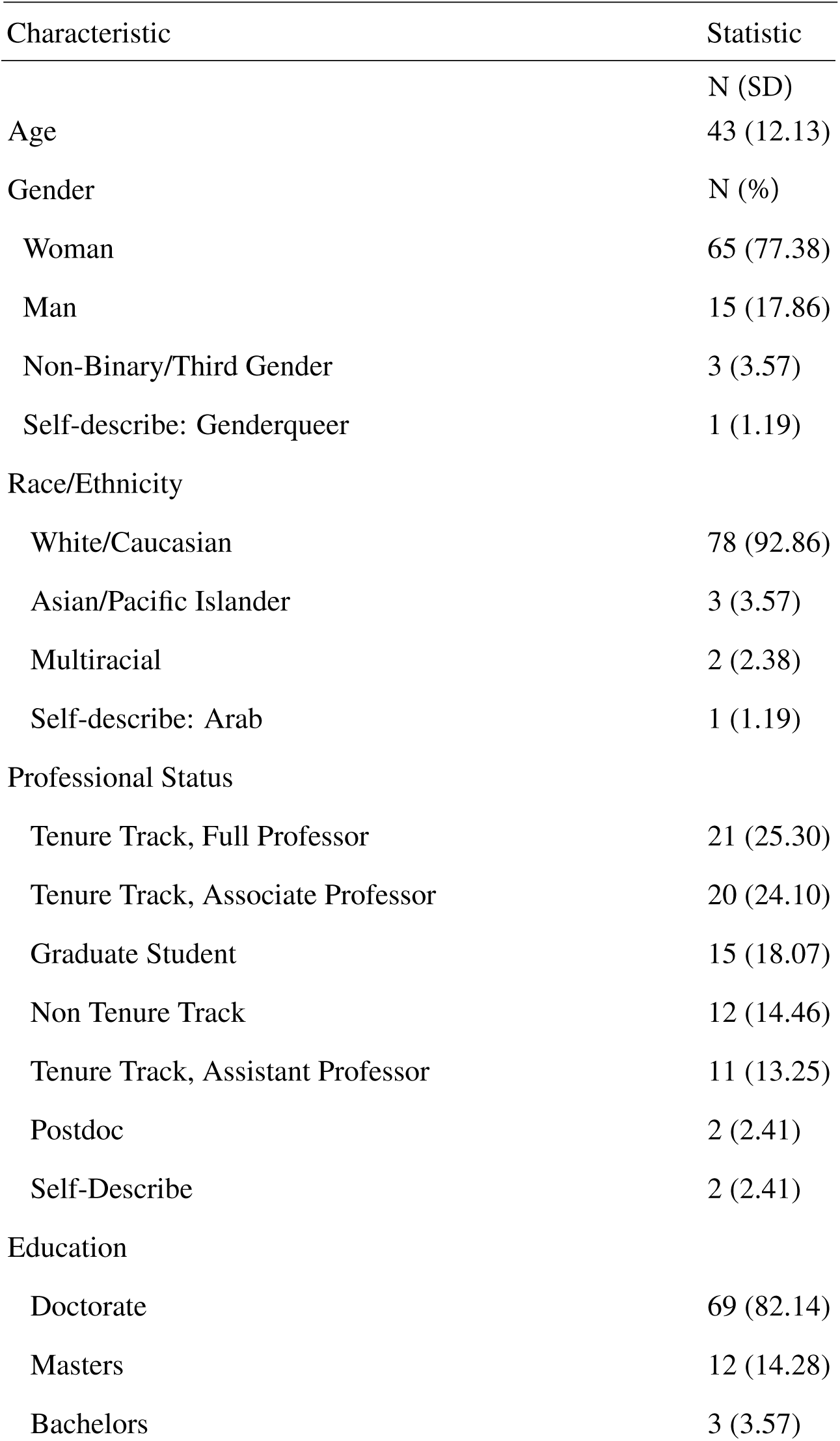
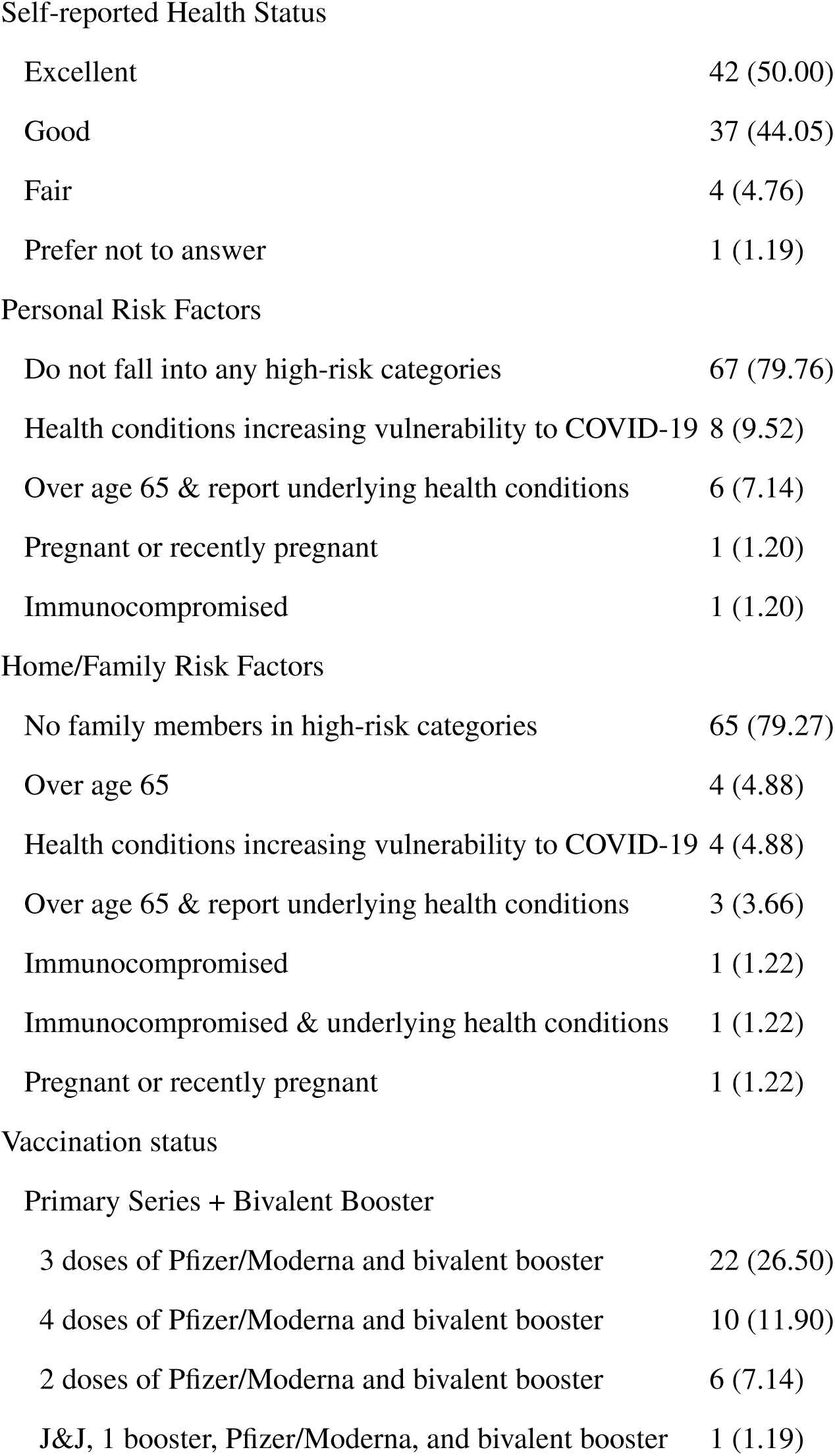
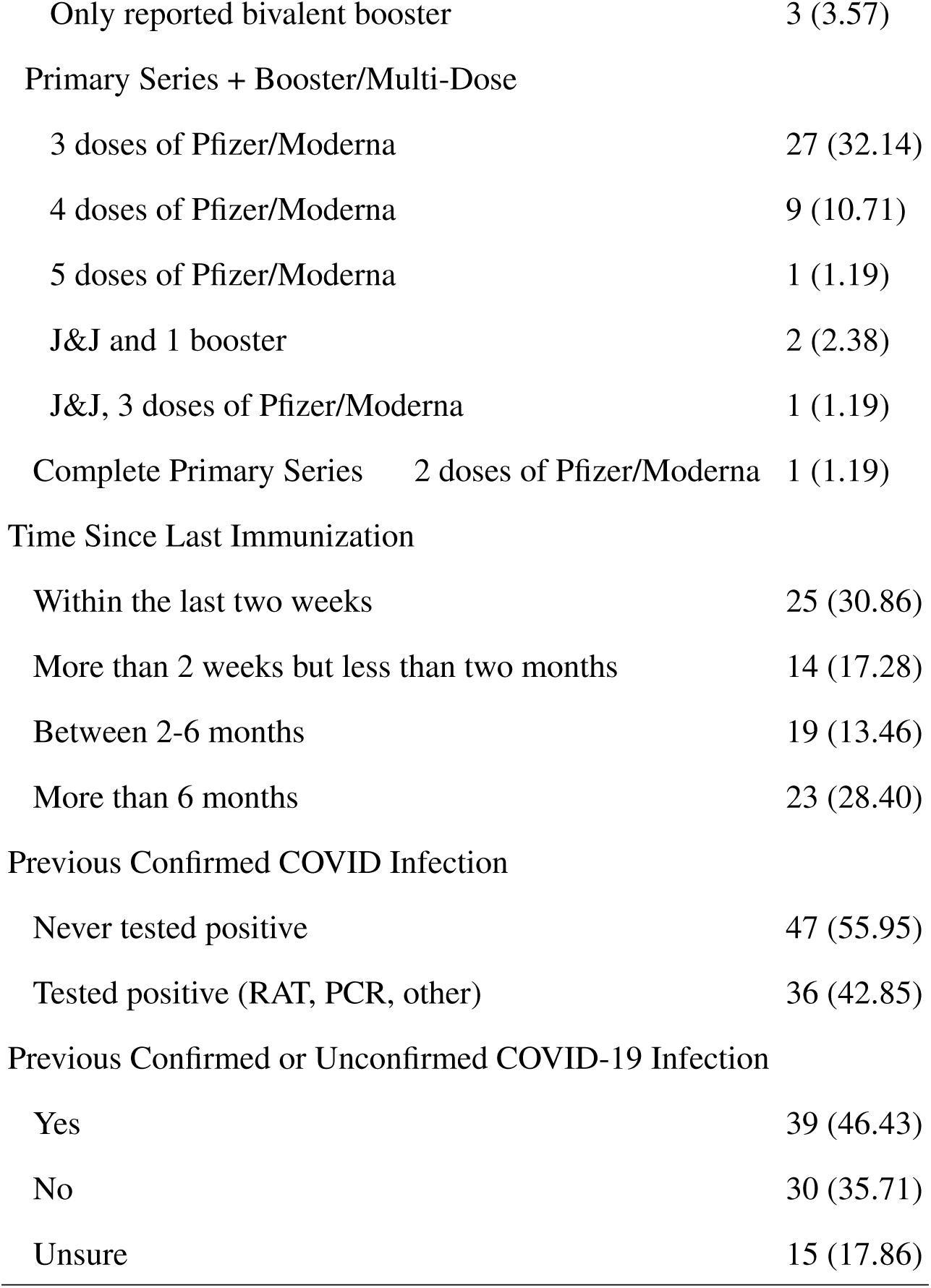
Participant demographics, professional status, personal and family health status. Underlying health conditions include those associated with increased risk of severe cases of COVID-19, including cardiovascular disease, diabetes, etc.)

Daily check-in surveys collected participants’ self-reported symptomatic status (based on CDC guidelines [2020], testing results from the current day, and reported mitigation behaviors from the previous day. For each check-in, participants were provided with a matrix of conference events from the previous day (panels, meals, workshops, etc.) and asked to report mitigation behaviors, including masking, handwashing, and distancing during each event. Symptomatic status and testing results were collected in post-conference check-ins. In addition, room dimensions, occupancy levels, and CO2 ppm for estimated air changes per hour (ACH) conversion were collected (See Supplement B). This study was approved by the Lehigh University Institutional Review Board, #1946144-1.

## Statistical Analysis

We define (i) ‘uptake’ as self-reported use of RATs before, during, and after the conference and as self-reported mask-wearing during the conference; (ii) ‘impact’ as self-reported positive test results from RATs, and (iii) ‘mitigation perceptions’ as respondents’ perception of the usefulness of specific mitigation measures implemented. RAT testing per day was measured as the proportion of attendees who answered ‘Yes’ when asked if they used an antigen test. Percent masking per day was measured dichotomously, based on whether participants reported masking during any of the events during each day of the conference. Continuous variables were summarized with means and standard deviations, and categorical variables are summarized with percent and frequency.

The association between RAT and masking uptake as dependent variables and the day in which the attendee was surveyed as the independent variable (pre-conference survey as reference) was modeled with logistic regression (Corresponding visual Fig. 2). Point estimates for odds ratio and 95% CI are reported as well as p-values. Logistic regression was used to estimate the association between RAT (and mask) uptake and predictors: daily symptomatic status, infection by COVID-19 before the conference, whether the participant has family members at increased risk of COVID-19, and whether the participant reports personal risk factors that increase of COVID-19. We used a generalized estimating equations (GEE) approach with a logistic binomial link function to model RAT use and masking as repeated-measures dependent variables. We used a compound symmetric correlation matrix. Results are presented as odds ratios, 95% CIs, and p-values (Table 3). RAT sensitivity for individual tests and the mean sensitivity were estimated by tracking (i) testing cadence, (ii) reported results, and (iii) symptomatic status. Sensitivity estimations for single, two-serial, and three-serial testing by symptomatic status are derived from the protocol in Soni et al. (2023). (See Supplement D for Testing cadences, reported results, and symptomatic statuses).

**FIG. 2.**
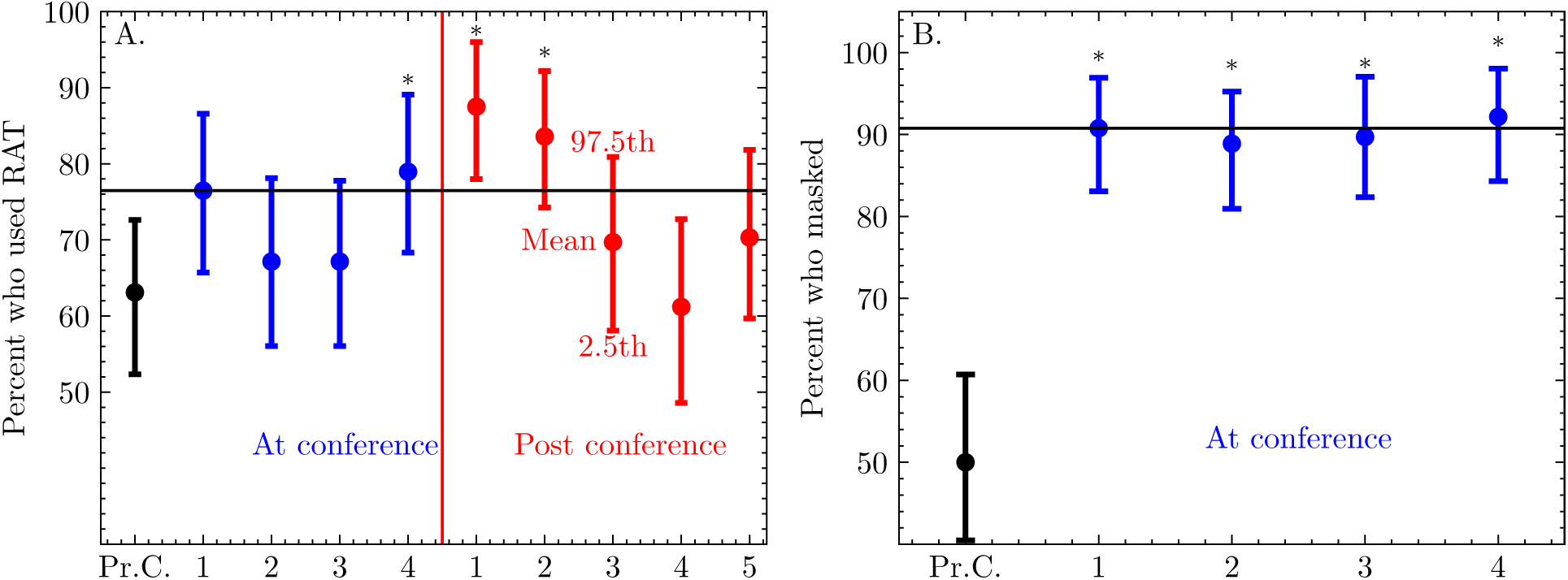
(A.) The percent of participants who reported using rapid antigen tests (RATs) before the conference started (black) (i.e., “Pre-conference” or Pr.C.), days 1-4 during the conference (blue), and one through 6 days after the conference ended (red). (B.) Percent of participants who reported masking at least once pre-conference (black) and during the conference (blue) which was filled out either on the last day of the conference or the day after the conference (relative to each person’s departure date). The mean is represented as a filled circle and the 95% confidence interval (using a Normal approximation) is represented as a vertical bar.

Statistical analyses were performed in STATA BE 17 and Python 3.

## Results

### Participants’ Backgrounds

Of the 258 in-person conference attendees, 84 (33%) participated in the pre-survey and at least one survey during or after the conference. Thirty-three participants completed all eleven surveys (Figure 3). The majority of participants identified as white (93%; 78/84), women (78%; 65/84), and having a doctorate degree (82%; 69/84; Table 1). The majority self-reported being in ‘Excellent’ health (50%; 42/84) and did not report any personal risk factors for COVID-19 (80%; 67/84). Most participants reported that family members did not suffer from personal risk factors (79%; 65/84). The mean age was 43 (Std. dev. 12.13), and a small proportion reported being over the age of 65 (7%; 6/84). Before attending the conference, 56% (47/84) reported never having a confirmed positive COVID-19 test, while only 46% (38/84) reported believing they had *never* contracted COVID outside of testing results. All participants reported being fully vaccinated, approximately half having received Moderna bivalent booster (mRNA-1273.214) before the conference (51%; 43/83).

**FIG. 3.**
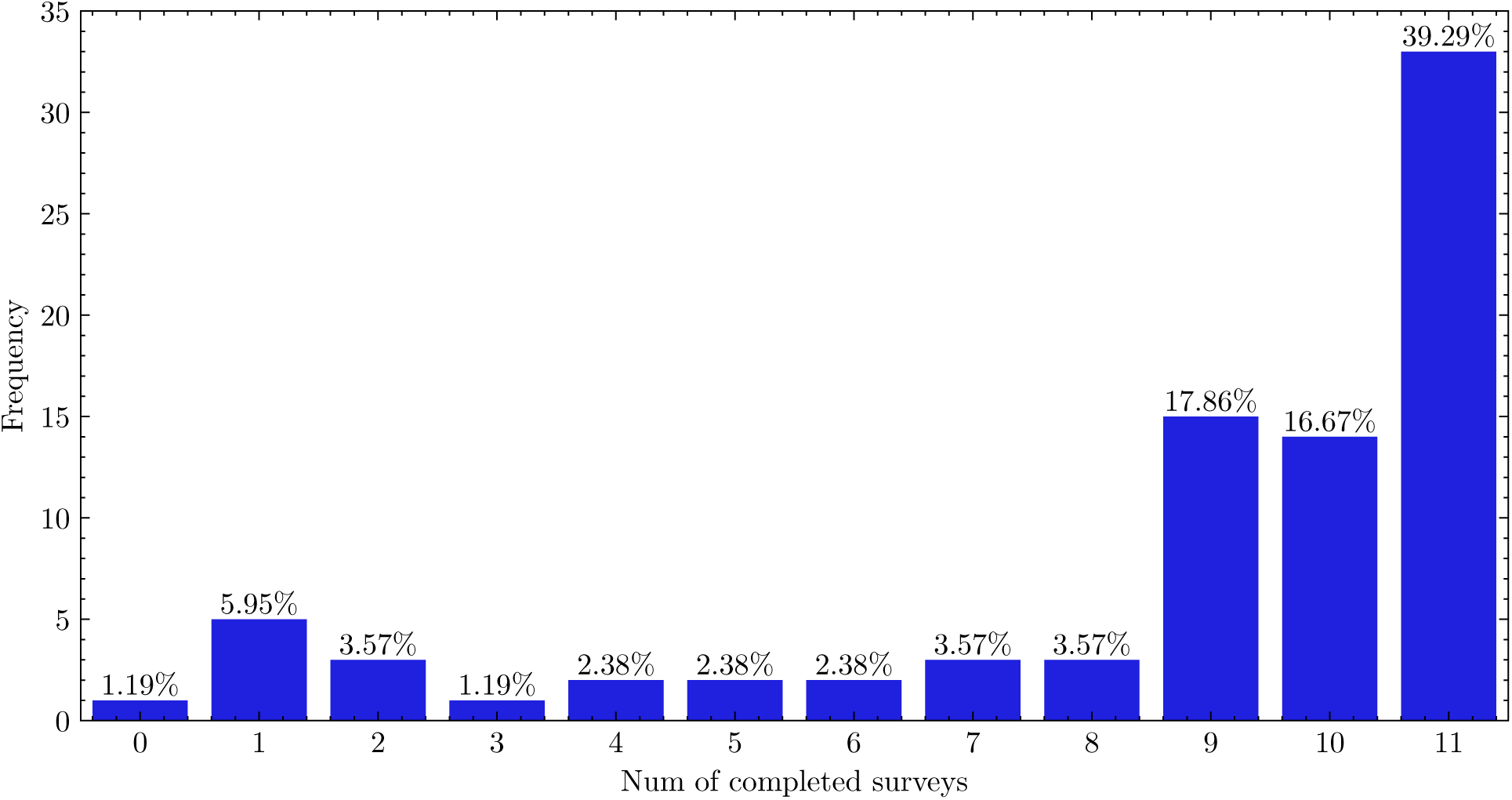
The number of participants who completed 0,1,2,…11 surveys over the observation period. The blue bars indicate frequency and percentages atop each bar are the number of completed surveys divided by 84, the number of participants who completed the pre-conference survey.

When provided a list of the mitigations the conference organizers implemented in the post-conference survey, participants indicated that they found access to free masks (91%; 58/64) and RATs (89%; 57/64; Table 2) the most helpful interventions. Participants felt that encouraging masks (97%; 62/64) and RATs (98%; 63/64) were the mitigations most helpful to others. The least helpful mitigation reported by participants for themselves and others was receiving local COVID-19 surveillance information (14%; 9/64).

**TABLE 2:** Attendee rating of most helpful mitigations that confer-ence organizers used target to the individual attendees (individual mitigations) and to the entire conference population and environ-ment (structural mitigations)

| Characteristic | Statistic |
| --- | --- |
| **Individual Mitigations** | N (%) |
| Access to | 58 (92.06) |
| Free N95 masks |  |
| Free rapid tests | 57 (89.06) |
| Basic public health information | 12 (18.75) |
| Local COVID-19 surveillance information | 9 (14.06) |
| **Structural Mitigations** | 63 (96.20) |
| *Encouraging* |  |
| Testing and making rapid tests freely accessible |  |
| Masking and making N95 masks freely accessible | 62 (95.38) |
| *Providing* | 12 (18.75) |
| Access to basic public health information |  |
| Local COVID-19 surveillance information | 9 (14.06) |
| Air ventilation/purification and measurement | 44 (67.69) |
| COVID relief funds/hybrid presentation option for those who<br>test positive before/during the conference | 35 (53.85) |

Overall, conference participants found the mitigation measures acceptable. In response to the post-survey question ‘Did any COVID mitigations seem unnecessary, intrusive, obtrusive, or counterproductive? If so, how?’ Most participants found mitigation measures ‘untroublesome’ (78%; 39/50); however, a few participants reported that masks inhibited social interaction (10%; 5/50). Discomfort with wearing masks was related to difficulty breathing (participant reporting asthma), recognizing people (participant reporting face blindness), hearing speech clearly, and encumbrance when donning and doffing masks.

### Uptake of RATs and Masks

Uptake of RATs and masks was highest on the last day of the conference and immediately post-conference (Figure 2). Compared to the proportion of participants who used RATs pre-conference (63%; 53/84), the proportion of participants using RATs was higher on the last day of the conference (79%; 45/57; *p*=0.05) and for two days after the conference ended (88%; 42/48; *p <*0.01 and 84%; 56/67; *p<*0.01)

RAT uptake was associated with reported symptomatic status (Table 3). Among participants who reported any testing during or after the conference, participants who reported experiencing one or more symptoms were also more likely to test (OR: 3.81; 95CI% [1.19, 7.63]; *p<*0.01). Previous infections of COVID-19, personal risk factors, and living with a family member who had one or more risk factors for severe outcomes from COVID-19 were not significantly associated with RAT use.

**TABLE 3.** Logistic regression using GEE and an exchangeable correlation matrix to estimate the association between masking and testing; and personal factors.

| Covariate | Odds Ratio | 95CI | p |
| --- | --- | --- | --- |
| **RAT Testing** |  |  |  |
| One or more Symptoms | 3.81 | [1.91 ,7.63] | < 0.01 |
| Any Personal Risk Factor | 2.90 | [0.63 ,13.24] | 0.17 |
| Any Family Risk Factor | 0.62 | [0.19, 2.07] | 0.44 |
| Prev. COVID Infection | 0.84 | [0.39, 1.83] | 0.67 |
| **Masking** |  |  |  |
| One or more Symptoms | 0.50 | [0.12, 2.04] | 0.32 |
| Any Personal Risk Factor | 4.72 | [0.67, 33.15] | 0.12 |
| Any Family Risk Factor | 1.29 | [0.22, 7.61] | 0.78 |
| Prev. COVID Infection | 0.96 | [0.31, 3.00] | 0.94 |

For masking, all conference days showed higher masking rates compared to the pre-conference baseline (Figure 2B). The odds of masking during the conference were double the odds reported before the conference, (OR: 2.00; 95% CI = [1.49, 2.70]; *p <*0.01; see Table 4). No predictors were significantly associated with masking during the conference (Table 3), though personal risk factors were the strongest predictor of mask use (OR: 4.72; 95CI% = [0.67, 33.15]; *p*=0.12).

### Conference Impact and Testing Results

Participants did not report any positive test results in the daily or post-conference check-in surveys. One month after the end of the conference, two participants reported positive cases of COVID-19 among the 258 conference registrants. One of these positive cases was reported to have occurred on day three of the conference and the second, seven days after the conference ended.

The mean sensitivity for all participants across the nine-day testing period was estimated to be 44.0% (95%CI = [37.1, 47.5]). Mean sensitivity peaked first day after the conference: Day 5 at 63.3% (95%CI = [54.8, 71.8]) (See Supplement D, Figures S3 and S4). Most participants tested daily, resulting in a mean difference between testing days of 23.1 hours (95% CI = [22.3, 23.9]).

## Discussion

The ‘COVID Conscious’ protocol implemented during the organization’s 2022 conference included multiple tools for reducing the risk of transmission of an infectious respiratory agent: (i) providing free rapid tests, (ii) providing high-quality N95 masks, (iii) providing basic public health safety information and local community risk profiles, (iv) reducing conference density, (v) monitoring ventilation and providing auxiliary air purification, and (vi) budgeting support for participants who had to cancel or remain isolated if they fell ill.

Our findings align with past work showing that multi-pronged approaches to prevent SARS-CoV-2 transmission during conferences tend to be accepted by attendees (Dupouy et al., 2023; Ehteshami et al., 2023; Gomes et al., 2022; Gottlieb et al., 2020; Silver et al., 2022; Sumer et al., 2022; Talcott et al., 2022). Two studies examined the uptake of multi-pronged mitigation strategies and positivity rates. Sumer et al. (2022) implemented a ‘safety concept’ model for the joint annual meeting of the Swiss Societies for Infectious Diseases (SSI) and Hospital Hygiene (SSHH) societies (September 2-4, 2020). The SSI/SSHH conference mandated masking, social distancing, and handwashing. Their cohort was unvaccinated and relied on self-reported PCR three weeks after the conference or seroconversion four weeks after the conference. The Hep-DART Conference (December 5-9, 2021) implemented a ‘Swiss Cheese’ model of multi-layered mitigation strategies during the initial acceleration of the first Omicron (BA.1) wave, which was similar to the NAVSA ‘COVID Conscious’ approach (Ehteshami et al., 2023). Consistent with our findings, both studies had a low reported number of cases (SSHI/SSI: no documented infections; Hep-DART, 2/52) and high rates of adherence to mitigations (SSHI/SSI, 80% surgical mask self-report; Hep-DART, no data). Although SSI/SSHH and Hep-DART mandated masking and (initial) testing, NAVSA measures were voluntary. A list of professional conferences across varied disciplines and their reported mitigation strategies can be found in Supplement F, Figure S5 (see Figure S6 for conference mitigation coding).

In reviewing professional conferences and the research on their mitigations, we found that structural mitigation of risk through monitoring and improving indoor air (IAQ) quality was lacking as an intervention. To our knowledge, no research has studied the effect or perceptions of monitoring and IAQ at academic conferences or similar events. As other work has noted, if present at all, attention to IAQ during the pandemic was sub-standard (Katz et al., 2023). While we were unable to assess the efficacy of our Corsi-Rosenthal boxes (See Appendix B, Figure S3 for sample room ACH data) continued experimental research suggests they are an effective way to reduce exposure to aerosol containing pathogens like SARS-CoV2 (Cadnum 2023). In our sample, respondents rated IAQ the third most helpful mitigation. Because this mitigation is an inexpensive structural intervention, requiring no behavior change on the part of individuals, it merits serious consideration as part of any risk reduction plan for events.

Beyond including IAQ, our study challenges the contention that the subject-matter expertise of conference attendees might factor into adherence to mitigations. Sumer et al. (2022) and Ehteshami et al. (2023) speculate that high adherence to conference mandates could have been due to participants’ expertise in infectious disease and medicine. The outbreak during the CDC’s EIS Conference (April 24–27, 2023) reported a low proportion of participants masking (CDC, 2023). In contrast, the NAVSA population—composed of humanists trained in literary studies and history—reported a high rate of masking and low-test positivity rates. Expertise in infectious diseases is neither necessary nor sufficient to engage in disease mitigation behaviors.

Subsequent work can address several limitations. Though this study reported uptake of masking and RATs, we cannot estimate to what degree specific mitigations reduced incident cases of COVID-19. We do not know if infections occurred during or after the conference or if positive tests went unreported. Selection bias may have attracted participants who had a pre-existing investment in COVID-19 mitigation measures. Social desirability may have contributed to over-reporting masking/RAT use and under-reporting infections.

The NAVSA sample’s uptake of mitigation behaviors may have been motivated by factors beyond personal risk reduction. Other measured motivators for masking and testing in the extant literature include a respect for others, ease of use, self-efficacy, and close contact during crowded events (Kwan et al., 2022; Lee et al., 2024). Given the sample’s overall health and vaccination status, participants’ overall steady masking rates may be indicative of these other motivators.

While participants masked at a steady rate, we found RAT uptake to be more variable. The highest frequency of testing occurred at the end and right after the conference (Fig. 2). Finally, as cost is an established barrier to testing, the availability of free RATs in the COVID Conscious Protocol may have incentivized more testing than participants would have normally performed.

The low sensitivity of RAT (Flowflex, the tests provided attendees: 67.4% overall for the Omicron variant compared to Delta) (Bayart et al., 2022) may have contributed to false negatives, especially considering recent estimates placing one-time asymptomatic testing sensitivity for BA.5 at 39.3% (Soni et al., 2023). Serial testing encouraged by the NAVSA COVID Conscious Protocol may have attenuated this limitation by improving RAT sensitivity to some degree (Chu et al., 2022). Recent research and updated recommendations have suggested that a two- and three-time testing cadence 48 hours apart increases the sensitivity of RATs up to 93% for symptomatic cases (2 tests, 48 hours apart) and up to 76% for asymptomatic cases (3 tests, 48 hours apart) (Soni et al., 2023). Applying this serial protocol to our sample helped provide an estimate of RAT uptake’s corresponding diagnostic performance: a mean sensitivity across the nine-day conference and post-conference testing of 44.0% (95%CI = [37.1, 47.5]) and a peak sensitivity of 63.3% (95%CI = [54.8, 71.8]), on day five.

This study’s testing uptake data and corresponding estimated diagnostic performance may provide a rough but potentially helpful benchmark for conference organizers, especially given that symptomatic status was associated with increased odds of RAT. A revised protocol for a four-day conference might encourage testing on any day when symptomatic and the first day, third day, and the day after the conference if asymptomatic to make the best use of tests, maximize sensitivity, and increase compliance. This protocol can also leverage a possible motivation for testing that may be inferred from our results. Future researchers and organizers should consider how data regarding motivation, cadence, and diagnostic accuracy can help facilitate an efficient screening component of a mitigation program.

Our sample of relatively healthy, fully vaccinated participants may have been at lower risk for severe COVID-19 outcomes. Just half of the conference attendees received the bivalent booster (51.8%, 43/83), which had become available to the general population three weeks before the start of the conference. This proportion was markedly higher than the US population 18 years of age or older during the same period (5.6% bivalent uptake, data from October 12, 2022) (CDC 2022b). Our sample’s vaccination and health status may have informed their high acceptance and uptake of mitigation measures (compared to their self-reported baselines during pre-conference daily and professional life). While personal risk factors and the presence of symptoms predicted increased odds of RAT uptake, no predictors were significantly associated with masking uptake.

Future work might evaluate the receptiveness to and uptake of mitigation measures under different temporal, geographical, contextual, and epidemiological parameters. The timeline of the COVID-19 pandemic should be considered when comparing behaviors from different conferences, as perceptions of risk, variants, and epidemiological burdens vary (Nguyen et al., 2022). Furthermore, data on participants’ circulation in community venues outside the conference might be parameterized with local surveillance data to model a conference’s ‘epidemiological footprint’ in more quantifiable terms.

Conference organizers may wish to consider the cost of mitigation measures and choose specific times to emphasize the use of masks and RATs. Because the uptake of masking was lower during the middle of the conference and the use of RATs was lowest mid-conference, organizers may wish to (i) reinforce the use of masking during the middle of a conference and (ii) suggest testing if attendees are symptomatic and at 48-hour serial testing cadence if asymptomatic, relative to conference duration for asymptomatic attendees.

### Conference Planning as Critical Public Health Practice

Beyond measuring uptake, perception, and impact, the results of this protocol invite an opportunity for critical public health practice. Events like conferences are not localized within planned space and times: they have wider social, cultural, and economic impacts (Shabee 2012). If professional organizations can become conscious of their events’ carbon footprints, we suggest they might, concordantly, become conscious of their event’s ‘epidemiological footprint.’

Accessibility to individual mitigation and conference space with structural mitigation measures may provide a more inclusive environment for individuals who are at high risk for complications from respiratory infections such as COVID-19. However, the benefits and potential harms should be considered in the decision-making process for event organization (Pearce et al. 2022). For instance, in our sample, those suffering from conditions such as face blindness and hearing impairment reported some encumbrance from others wearing masks, a problem that was compounded by the noise produced by Cori-Rosenthal purifiers. Hybrid conferencing has been an oft-proposed measure to increase accessibility, especially for those with disabilities, including those who are at high risk for complications from COVID-19. While hybrid options can compound costs and logistical complexity, recent studies found that hybrid options expand inclusivity but can also entail accessibility barriers (Alharbi et al., 2023).

Cost is a well-established barrier to attending professional conferences in academia (Velin et al., 2021). The ‘COVID Conscious’ protocol proved relatively cost-efficient and did not increase registration fees, an important consideration for academic conference organizers wishing to lower the financial barriers for attendance. The total cost of mitigation measures and relief aid was approximately $6,482, or $25 per conference attendee (Supplement E, Table S1). The financing of NAVSA’s ‘COVID Conscious’ approach could be scaled to larger conferences by adding mitigation fees to conference costs or by inviting a sponsor to subsidize. Future organizers might consider a more conservative allocation of RATs following a serial testing protocol, further reducing costs. The conference organizers reported two refunds requested due to illness, and one reported relief request for leaving the conference early due to illness, for a total of $1,067.40 (17% of the mitigation budget). Allocation and distribution of such relief may pose a challenge when scaled to larger conferences.

Reflecting and acting critically on public health implications and opportunities of professional events requires organizers to consider how a conference can converge various “publics” in space and time: that the population affected and affecting larger public health extends beyond academics and their specific structural inequities. The occupational health of venue employees is an important consideration, given the existing disparities workers in this industry experience and whose risks of infection and job precarity were magnified during the earlier waves of the COVID-19 pandemic (Rosemberg et al., 2021; Yan et al., 2022). If the trend in various academic organizations’ policies to prefer or exclusively contract unionized hotels--to support more equitable working conditions for hotel employees--continues,^1^ we suggest that actively mitigating a conference’s ‘epidemiological footprint’ for hospitality workers would be a congruous goal.

This concern invites more expansive consideration of how conferences affect public health in their host cities. To estimate the impact of the conference on the surrounding community, studies could collect where and how frequently conference attendees contact the community, as well as the mitigations that attendees follow during these events. Depending on the incidence and risk of respiratory infections of concern, additional conferences could be studied under similar mitigation protocols and surveys to estimate the uptake of masks/RATs and how interactions between the conference and community may drive transmission.

Some examples include but are not limited to: American Public Health Association, American Studies Association, American Sociological Association, Modern Language Association, American Philosophical Association, American Historical Society, Organization of American Historians, International Studies Association

## Conclusion

The uptake of masks/RATs and a low number of positive infections at the NAVSA conference suggest that conference-level mitigations may have positively impacted the health of the conference population, the venue hospitality workers and guests, and the local communities within Northampton County. A multi-layered mitigation plan (Appendix A-E) can be financially feasible and conducive to inclusivity and accessibility for conference attendees. Because conference events connect the conference population with the local community, we suggest that conference-level mitigation can act as a public health intervention by potentially reducing the infectious disease burden conferences might introduce in terms of local population and occupational health. The magnitude of this effect will depend on a community’s epidemiological characteristics and an infectious agent’s transmission dynamics. The ‘COVID Conscious’ approach can be a helpful tool for professional organizations to plan safer conferencing for their membership and host communities in the face of novel epidemics and seasonal respiratory infectious diseases.

## Ethics Statement

The Lehigh University Institutional Review Board Lehigh approved this study with an exempt determination #1946144-1 on 9/17/2022. The Lehigh IRB granted an except determination because all of the participants were over the age of 18 and one or more of the following criteria were met: the information obtained is recorded by the investigator in such a manner that 1) the identity of the human subjects cannot be readily ascertained, directly or through identifiers linked to the subjects; 2) any disclosure of the human subjects’ responses outside the research would not reasonably place 3) the subjects at risk of criminal or civil liability or be damaging to the subjects’ financial standing, employability, educational advancement, or reputation; or, 4) the information obtained is recorded by the investigator in such a manner that the identity of the human subjects can be readily ascertained, directly or through identifiers linked to the subjects, and the IRB conducted a limited review to determine that there are adequate provisions to protect the privacy of subjects and to maintain the confidentiality of data Informed consent was obtained from the participants in this study. In addition, confidentiality of participants involved was assured throughout the data collection and all methods were carried out in accordance with relevant guidelines and regulation.

## Author Contributions

LS had the main responsibility for conceptualization, methodology, data collection, project administration, data analysis, and writing the manuscript. TM performed statistical analysis. TM made substantial contributions to the conceptualization, methodology, analysis of the data, and writing, and revision of the manuscript. KB made substantial contributions to the conceptualization, methodology, analysis of the data, and writing, and revision of the manuscript. ML made substantial contributions to the conceptualization, methodology, data collection, analysis of the data, and writing and revision of the manuscript. CC made substantial contributions to the conceptualization, methodology, analysis of the data, and writing and revision of the manuscript. JKM made substantial contributions to the conceptualization, methodology, analysis of the data, and writing and revision of the manuscript. AS made substantial contributions to the conceptualization, methodology, collection of the data and revision of the manuscript. LK made substantial contributions to the analysis of the data, and writing, and revision of the manuscript. BE made substantial contributions to the data collection, analysis of the data, writing and revision of the manuscript. All authors read the final version of manuscript for publication and agreed to be responsible for all aspect of the manuscript that in ensuring that questions related to accuracy or integrity of any part of the manuscript are appropriately investigated and resolved. All authors approved the final version to be published.

## Supporting information

Appendix A: COVID Conscious material

Appendix B: Air ventilation auditing and auxiliary purification

Appendix C: Survey Instruments

Appendix D: Sensitivity Estimates

Appendix E: Budget

## Acknowledgments

We would like to acknowledge two undergraduate students at Lehigh University, Will Yeager (IDEAS) and Tess Jones (Earth and Environmental Sciences), for their help in the design, building, and deployment of Corsi-Rosenthal boxes. We would also like to thank the group of undergraduate student volunteers from the Health, Medicine, and Society Program in the College of Arts and Sciences, the Community, Global, and Population Health in the College of Health at Lehigh University that helped with counting occupancy during the conference.

We would like to express our gratitude for the Health, Medicine, and Society Program and the College of Arts and Sciences at Lehigh University for helping fund the mitigation measures at the conference. We also want to thank ARANET for the discounted rate on CO2 meters.

## Data Availability Statement

All data that were used for analysis is available on a private Github repository. Interested readers may contact the first and corresponding author, Lorenzo Servitje, for access.

## Abbreviations

ACH: Air Exchanges Per Hour
EIS: Epidemic Intelligence Service
IAQ: Indoor Air Quality
NAVSA: North American Victorian Studies Association
RAT: Rapid Antigen Tests
SSI/SSHH: Swiss Societies for Infectious Diseases (SSI) and Hospital Hygiene (SSHH)

## References

1. Aldekheel, M., Altuwayjiri, A., Tohidi, R., Jalali Farahani, V., & Sioutas, C. (2022). The role of portable air purifiers and effective ventilation in improving indoor air quality in university classrooms. International Journal of Environmental Research and Public Health, 19(21), 14558.

2. Alharbi, R., Tang, J., & Henderson, K. (2023). Accessibility barriers, conflicts, and repairs: Understanding the experience of professionals with disabilities in hybrid meetings. Proceedings of the 2023 CHI Conference on Human Factors in Computing Systems, 1–15. 10.1145/3544548.3581541

3. Allen, J. G. & Ibrahim, A. M. (2021). Indoor air changes and potential implications for SARS-CoV-2 transmission. Journal of the American Medical Association, 325(20), 2112–2113.

4. Bakó-Biró, Z., Clements-Croome, D. J., Kochhar, N., Awbi, H. B., & Williams, M. J. (2012). Ventilation rates in schools and pupils’ performance. Building and Environment, 48(FEV), 215–223. 10.1016/j.buildenv.2011.08.018

5. Batterman, S. (2017). Review and extension of CO2-based methods to determine ventilation rates with application to school classrooms. International Journal of Environmental Research and Public Health, 14(2), 145.

6. Bayart, J.-L., Degosserie, J., Favresse, J., Gillot, C., Didembourg, M., Djokoto, H. P., Verbelen, V., Roussel, G., Maschietto, C., Mullier, F., Dogné, J.-M., & Douxfils, J. (2022). Analytical sensitivity of six SARS-CoV-2 rapid antigen tests for Omicron versus Delta variant. Viruses, 14(4), 654-. 10.3390/v14040654.

7. Bell, K. & Green, J. (2015). Keeping a critical edge: Reflections on 25 years as a scholarly journal. Critical Public Health, 25(1), 1–3.

8. Brook, C. E., Northrup, G. R., Ehrenberg, A. J., IGI SARS-CoV Testing Consortium, Doudna, J. A., & Boots, M. (2021). Optimizing COVID-19 control with asymptomatic surveillance testing in a university environment. Epidemics, 37, 100527.

9. Cadnum, J. L., Bolomey, A., Jencson, A. L., Wilson, B. M., & Donskey, C. J. (2023). Effectiveness of commercial portable air cleaners and a do-it-yourself minimum efficiency reporting value (MERV)-13 filter box fan air cleaner in reducing aerosolized bacteriophage MS2. Infection Control & Hospital Epidemiology, 44(4), 663–665

10. Centers for Disease Control and Prevention. (2020). COVID-19 and your health. https://www.cdc.gov/coronavirus/2019-ncov/vaccines/recommendations.html

11. Centers for Disease Control and Prevention. (2021). Ventilation in schools and childcare programs. https://www.cdc.gov/coronavirus/2019-ncov/community/schools-childcare/ventilation.html

12. Centers for Disease Control and Prevention. (2022). Covid data tracker weekly review. https://covid.cdc.gov/covid-data-tracker/#datatracker-home

13. Centers for Disease Control and Prevention. (2022) COVID-19 Vaccinations in the United States, Jurisdiction. https://data.cdc.gov/Vaccinations/COVID-19-Vaccinations-in-the-United-States-Jurisdi/unsk-b7fc/about_data

14. Centers for Disease Control and Prevention. (2023). *Covid by county.* https://archive.cdc.gov/#/details?url= https://www.cdc.gov/coronavirus/2019-ncov/your-health/covid-by-county.html

15. Centers for Disease Control and Prevention. (2023, May 26). Update on rapid assessment of SARS-CoV-2 transmission at 2023 EIS conference. https://www.cdc.gov/media/releases/2023/s0526-eis.html.

16. Christie, A., Brooks, J. T., Hicks, L. A., Sauber-Schatz, E. K., Yoder, J. S., Honein, M. A., COVID, C., & Team, R. (2021). Guidance for implementing COVID-19 prevention strategies in the context of varying community transmission levels and vaccination coverage. Morbidity and Mortality Weekly Report, 70(30), 1044.

17. Chu, V. T., Schwartz, N. G., Donnelly, M. A. P., Chuey, M. R., Soto, R., Yousaf, A. R., Schmitt-Matzen, E. N., Sleweon, S., Ruffin, J., Thornburg, N., Harcourt, J. L., Tamin, A., Kim, G., Folster, J. M., Hughes, L. J., Tong, S., Stringer, G., Albanese, B. A., Totten, S. E., … Matanock, A. (2022). Comparison of home antigen testing with RT-PCR and viral culture during the course of SARS-CoV-2 infection. Archives of Internal Medicine, 182(7), 701–709. 10.1001/jamainternmed.2022.1827

18. Di Gilio, A., Palmisani, J., Pulimeno, M., Cerino, F., Cacace, M., Miani, A., & de Gennaro, G. (2021). CO2 concentration monitoring inside educational buildings as a strategic tool to reduce the risk of SARS-CoV-2 airborne transmission. Environmental Research, 202, 111560.

19. Dupouy, J., Chaneliere, M., Schuers, M., Laporte, C., Bayen, M., Gaultier, A., & Rat, C. (2023). A face-to-face national congress experience during the COVID-19 pandemic: A report focusing on the risk of COVID-19 contamination. European Journal of General Practice, 29(2), 2139825.

20. Ehteshami, M., Edgar, C. L., Delgado Ayala, L. Y., Hagan, M., Martin, G. S., Lam, W., & Schinazi, R. F. (2023). Lessons learned from in-person conferences in the times of COVID-19. International Journal of Environmental Research and Public Health, 20(1), 510.

21. Food and Drug Administration. (2022, August 31). Coronavirus (COVID-19) update: FDA authorizes Moderna, Pfizer-BioNTech bivalent COVID-19 vaccines for use as a booster dose. Press release.

22. Goldberg, D. S. (2021). Against the medicalization of public health (ethics). Public Health Ethics, 14(2), 117–119.

23. Gomes, C. M., Souza, J. D. d., Anzolch, K. M., Henriques, J. V. T., Nogueira, L., Pimentel, E., Fernandes, R. d. C., Canalini, A. F., & Bessa Jr, J. D. (2022). Is it safe to resume large scale in-person medical meetings? International Brazilian Journal of Urology, 48, 857–863.

24. Gottlieb, M., Landry, A., Egan, D. J., Shappell, E., Bailitz, J., Horowitz, R., & Fix, M. (2020). Rethinking residency conferences in the era of COVID-19. AEM Education and Training, 4(3), 313–317.

25. Gouge, D. H., Lame, M. L., Stock, T. W., Rose, L. F., Hurley, J. A., Lerman, D. L., Nair, S., Nelson, M. A., Gangloff-Kaufmann, J., McSherry, L., Connett, J. F., Graham, L., & Green, T. A. (2023). Improving environmental health in schools. Current Problems in Pediatric and Adolescent Health Care, 53(4), 101407–101407. 10.1016/j.cppeds.2023.101407

26. Green, J., Fischer, E. F., Fitzgerald, D., Harvey, T. S., & Thomas, F. (2022). The publics of public health: Learning from COVID-19. Critical Public Health, 32(5), 592–599.

27. He, R., Liu, W., Elson, J., Vogt, R., Maranville, C., & Hong, J. (2021). Airborne transmission of COVID-19 and mitigation using box fan air cleaners in a poorly ventilated classroom. Physics of Fluids, 33(5).

28. Huang, J., Fisher, B. T., Tam, V., Wang, Z., Song, L., Shi, J., La Rochelle, C., Wang, X., Morris, J. S., Coffin, S. E., & Rubin, D. M. (2022). The effectiveness of government masking mandates on COVID-19 county-level case incidence across the United States, 2020: Study examines the effectiveness of US government masking mandates during a portion of the COVID-19 pandemic. Health Affairs, 41(3), 445–453. 10.1377/hlthaff.2021.01072

29. Katz, A., Li, T., James, L., Siegel, J., & O’Campo, P. (2023). Systematically omitting indoor air quality: Sub-standard guidance for shelters, group homes and long-term care in Ontario during the COVID-19 pandemic. Critical Public Health, 33(5), 683–696. 10.1080/09581596.2023.2262736

30. Kwan, T. H., Wong, N. S., Chan, C. P., Yeoh, E. K., Wong, S. Y.-s., & Lee, S. S. (2022). Mass screening of SARS-CoV-2 with rapid antigen tests in a receding omicron wave: Population-based survey for epidemiologic evaluation. JMIR Public Health and Surveillance, 8(11), e40175–e40175. 10.2196/40175

31. Lee, G. Y. L. & Lim, R. B. T. (2024). Are self-test kits still relevant post COVID-19 pandemic? Qualitative study on working adults’ perceptions. *Infection*, Disease & Health, 29(2), 73–80.

32. Lipsitch, M. & Dean, N. E. (2020). Understanding COVID-19 vaccine efficacy. Science, 370(6518), 763–765.

33. McNeill, V. F., Corsi, R., Huffman, J. A., King, C., Klein, R., Lamore, M., Maeng, D. Y., Miller, S. L., Ng, N. L., Olsiewski, P., Godri Pollitt, K. J., Segalman, R., Sessions, A., Squires, T., & Westgate, S. (2022). Room-level ventilation in schools and universities. *Atmospheric Environment*, X, 13, 100152, 10.1016/j.aeaoa.2022.100152

34. Morawska, L., Tang, J. W., Bahnfleth, W., Bluyssen, P. M., Boerstra, A., Buonanno, G., Cao, J., Dancer, S., Floto, A., Franchimon, F., Haworth, C., Hogeling, J., Isaxon, C., Jimenez, J. L., Kurnitski, J., Li, Y., Loomans, M., Marks, G., Marr, L. C., … Yao, M. (2020). How can airborne transmission of COVID-19 indoors be minimised? Environment International, 142, 105832. 10.1016/j.envint.2020.105832

35. NAVSA. (2022). NAVSA: Policies & procedures. https://navsa2022.org/covid-conscious/policies-procedures/.

36. Nguyen, N., Lane, B., Lee, S., Gorman, S. L., Wu, Y., Li, A., Lu, H., Elhadad, N., Yin, M., & Meyers, K. (2022). A mixed methods study evaluating acceptability of a daily COVID-19 testing regimen with a mobile-app connected, at-home, rapid antigen test: Implications for current and future pandemics. Plos one, 17(8), e0267766.

37. Pearce, E., Kamenov, K., Barrett, D., & Cieza, A. (2022). Promoting equity in health emergencies through health systems strengthening: Lessons learned from disability inclusion in the COVID-19 pandemic. International Journal for Equity in Health, 21(3), 149.

38. Persily, A. & de Jonge, L. (2017). Carbon dioxide generation rates for building occupants. Indoor air, 27(5), 868–879.

39. Shabajee, P., Hiom, D., & Priest, C. (2012). Rethinking Events in Higher Education: A Systemtic Sustainability Perspective. In Pernecky, T., & Lück, M. (Eds.). Events, society and sustainability : Critical and contemporary approache. (pp 58–76). Taylor & Francis Group

40. Rosemberg, M.-A. S., Adams, M., Polick, C., Li, W. V., Dang, J., & Tsai, J. H.-C. (2021). COVID-19 and mental health of food retail, food service, and hospitality workers. Journal of Occupational and Environmental Hygiene, 18(4-5), 169–179.

41. Schrecker, T. (2022). What is critical about critical public health? Focus on health inequalities. Critical Public Health, 32(2), 139–144. 10.1080/09581596.2021.1905776

42. Silver, C. M., Joung, R. H., Visenio, M. R., Wang, T. S., Pawlik, T. M., Kim, E. S., & Bilimoria, K. Y. (2022). COVID-19 positivity following an in-person surgical society meeting: A cross-sectional survey study. Journal of Surgical Research, 278, 267–270.

43. Soni, A., Herbert, C., Lin, H., Yan, Y., Pretz, C., Stamegna, P., Wang, B., Orwig, T., Wright, C., Tarrant, S., Behar, S., Suvarna, T., Schrader, S., Harman, E., Nowak, C., Kheterpal, V., Rao, L. V., Cashman, L., Orvek, E., … McManus, D. D. (2023). Performance of rapid antigen tests to detect symptomatic and asymptomatic SARS-CoV-2 infection: A prospective cohort study. Annals of Internal Medicine, 176(7), 975–982. 10.7326/M23-0385

44. Stokel-Walker, C. (2022). What do we know about COVID vaccines and preventing transmission? BMJ, 376.

45. Swaminathan, A., Smith, J, & Choo, E. (2022). The irony — and ignominy — of medical conferences as superspreader events. StatNews. https://www.statnews.com/2022/06/14/the-irony-and-ignominy-of-medical-conferences-as-superspreader-events/

46. Suh, H. H., Meehan, J., Blaisdell, L., & Browne, L. (2022). Non-pharmaceutical interventions and COVID-19 cases in us summer camps: Results from an American Camp Association survey. Journal of Epidemiology and Community Health, 76(4), 327–334. 10.1136/jech-2021-216711

47. Sumer, J., Flury, D., Kahlert, C. R., Mueller, N. J., Risch, L., Nigg, S., Seneghini, M., Vernazza, P., Schlegel, M., & Kohler, P. (2022). Safety evaluation of a medical congress held during the COVID-19 pandemic—A prospective cohort. International Journal of Public Health, 67, 1604147. 10.3389/ijph.2022.1604147

48. Talcott, W. J., Chen, K., Peters, G. W., Reddy, K. K., Weintraub, S. M., Mougalian, S. S., Adelson, K., & Evans, S. B. (2022). Self-reported COVID-19 infections and social mixing behavior at oncology meetings. International Journal of Radiation Oncology*Biology*Physics, 114(1), 30–38.

49. Talic, S., Shah, S., Wild, H., Gasevic, D., Maharaj, A., Ademi, Z., Li, X., Xu, W., Mesa-Eguiagaray, I., Rostron, J., Theodoratou, E., Zhang, X., Motee, A., Liew, D., & Ilic, D. (2021). The effectiveness of non-pharmaceutical interventions in reducing SARS-CoV-2 transmission and COVID-19 incidence and mortality: systematic review and meta-analysis. British Medical Journal (BMJ), 2021(375), e068302.10.1136/bmj-2021-068302

50. Velin, L., Lartigue, J.-W., Johnson, S. A., Zorigtbaatar, A., Kanmounye, U. S., Truche, P., & Joseph, M. N. (2021). Conference equity in global health: A systematic review of factors impacting LMIC representation at global health conferences. BMJ Global Health, 6(1), e003455. 10.1136/bmjgh-2020-003455

51. Yan, J., Kim, S., Zhang, S. X., Foo, M.-D., Alvarez-Risco, A., Del-Aguila-Arcentales, S., & Yáñez, J. A. (2021). Hospitality workers’ COVID-19 risk perception and depression: A contingent model based on transactional theory of stress model. International Journal of Hospitality Management, 95, 102935.

