## Appendix A: COVID Conscious material for "Can a Conference be Epidemiologically Conscious? A Pilot Study of Implementing COVID-19 Mitigation Measures at an In-Person Conference"

The following supplement contains an archive of the public-facing NAVSA 2022 “COVID Conscious” material that were made available to participants on the NAVSA2022.org website and were communicated by email, social media, and conference mobile app (Ex Ordo) before and during the conference. This material remains available on the navsa2022.org website.

Conference Policies and Procedures Page [nav]

We want to assure you that we are committed to providing a safe environment for all. We will continue to monitor local requirements as we get closer to the event date and update our protocols as new information becomes available.

We will adhere to COVID-19 safety guidelines established by the U.S. Centers for Disease Control and Prevention, local health authorities, and the conference venue. We will share any special protocols at our event with you as the date approaches.

In accordance with local guidelines, masks are currently recommended for all participants in public spaces, regardless of vaccination status.

We ask anyone feeling ill or showing symptoms of COVID-19 to remain home. If you have had contact with or been exposed to anyone who has tested positive for COVID-19 in the 14 days prior to the event, we ask that you test and monitor your symptoms before attending.

If you are unable to participate for a COVID-19-related reason, please let us know immediately, and we will work with you on your registration refunds or see about potential remote accommodations.

<u>Before the conference</u>

Please familiarize yourself with current CDC recommendations regarding travel and local COVID-19 requirements. While masking is not currently mandated while traveling, masing during travel reduces your chance of exposure (especially when planes are unventilated before take-off and on landing)

Understand that public spaces have an inherent risk of exposure to COVID-19 and evaluate your comfort level with attending an in-person conference.

Please stay home if you are not feeling well or have had COVID-19 exposure in the 14 days before the event. Contact the organizers if this applies to you.

We encourage everyone to test before traveling to the event. If you would like a free antigen test, please visit this link.

During the conference

We encourage everyone to wear well-fitted masks in the conference space unless you are actively eating.

Maintain appropriate social distancing at high-traffic areas such as the registration desk and respect posted social distancing signs.

Recognize some conference events (meals and receptions) where attendees will be unmasked while eating/drinking. Consider your comfort level when deciding to participate in these events.

Follow all appropriate recommendations, including frequent hand washing, covering your mouth and nose while sneezing or coughing, and avoiding face touching. To assist in minimizing potential physical contact, elbow bumps are a great alternative to handshakes.

Should you feel ill or exhibit COVID-19 symptoms while attending the conference, we ask that you quarantine in your room and contact the conference coordinators at or (833) 676-3967 for help with any support you might need.

After the conference

If you test positive for COVID-19 within 10 days of returning home, we encourage you to contact the conference coordinators at or (833) 676-3967 and notify other conference participants that you were around.

**Getting Help Page**

Getting Help

What to do if you test positive or feel sick? (Link to CDC)

Though JV organizers can't provide medical advice, we are available to assist conference participants in answering questions, provide masks and tests free of charge, and help address concerns about logistics in the event of illness. Please contact us if you cannot attend the conference due to illness. We can provide you with a refund or can try to find a presentation accommodation to the best of our ability.

We encourage attendees to test during the conference and also ask that attendees not attend events if they are positive. If you have a positive result on a rapid test and need help getting a confirmatory rapid-PCR test or any other support, please text (833) 676-3967. Support can include: needing help with PPE (personal protective equipment), tests, food delivery, and/or financial and logistical assistance getting/paying for a single room if you are sharing.

```
Urgent Care Centers near Conference Venues
Patient First Primary and Urgent Care
2310 Schoenersville Road,
Bethlehem, PA 18017-3602
(484) 403-7560
8 am to 10 pm, Daily
No appointment necessary

St. Luke's Care Now { Bethlehem (Walk-in care) & Occupational Medicine
153 Brodhead Road Bethlehem,
PA 18017, (484) 526-3000
```

8 am to 9 pm, Monday-Friday
8 am to 9 pm, Saturday/Sunday
No appointment necessary

### Emergency Rooms

St. Lukes Hospital
801 Ostrum St. Bethlehem,
PA 18015, (866) 785-8537

Lehigh Valley Hospital { Muhlenberg
2545 Schoenersville Road Bethlehem,
PA 18017, (484) 884-2521

### Local Test-to-Treat Sites

CVS Store #01036
2434 Catasauqua Road Bethlehem,
PA 18018 (610) 868-5122
Book an appointment at CVS Store #01036

CVS Store #01093
7001 North Highway, 309 Coopersburg,
PA 18036, (610) 282-4104
Book an appointment at CVS Store #01093

CVS Store #02296
3295 PA Route 100 Macungie,

PA 18062, (610) 967-5684

Book an appointment at CVS Store #02296

Public Health Links

- CDC Guidelines Regarding Symptoms, Testing, and Treatment
- Pennsylvania “Stop the Spread”
- Information on Monkey Pox

**Prevention Page**

Masking

Masks are strongly encouraged throughout the conference. 3M, Aura, N95 masks, (and surgical masks), and antigen tests will be available at registration and the welcome desks daily.

Please text (833) 676-3967 if you need a mask, test, or other support.

We are doing our best logistically to reduce crowding, assess real-time ventilation estimates, and place our indoor banquet in a room that is sized for double occupancy and has extraordinarily high ceilings.

Risk Measures

Below are two metrics used by communities to assess the degree of risk concerning COVID-19: Community Transmission Levels in Northampton County, PA <Applet created using COVID NOW DataAPI>

What is community transmission?

This metric measures the level of confirmed cases locally per 100,000 and assigns a risk level. “High” transmission is anything greater than 50 cases per 100,000 or a positivity rate greater than 10%. The positivity rate is the number of positive tests / the number of tests reported.

### How to interpret community transmission

This depends on who you ask: some infectious disease experts feel comfortable unmasking in indoor spaces when cases are somewhere between 1 and 10 per 100,000. Along with the trend or direction of community transmission (going up or down over the last week), the positivity rate can help determine if the community transmission cases are an undercount. In the current COVID climate, positivity rates are substantially high (unsurprisingly because of the amount of testing done and not reported, as in-home tests), so community transmission can be thought of as a floor." Unless positivity rates are low (¡10%), as a guideline, community transmission (cases per 100,000) represents a minimum level of community transmission.

### What is the community transmission in my home location?

<Applet created using COVID NOW DataAPI>

### Community Risk Levels in Northampton County, PA

<Applet created using COVID NOW DataAPI>

### What are community risk levels?

Community Risk is a "new tool to help communities decide what prevention steps to take based on the latest data." It is derived from the number of available hospital beds, the number of hospital beds used, and the number of new local cases. The CDC changed its primary reporting mode from Community Transmission to "Community Risk" on February 25.

We provide this metric given that it is the one communicated by CDC, but would suggest that Community Risk Levels is less useful on top of being a lagging measure. Community Risk Levels are a contested measure amongst the public health and epidemiological communities.

Given the variables and thresholds involved, Community Risk Level is more of a health system measure, reflecting the capacity of a community to manage severe cases in hospitals. For individuals, this might translate to the risk of having severe covid and not being able to get treatment.
