## Appendix B: Air ventilation auditing and auxiliary purification for "Can a Conference be Epidemiologically Conscious? A Pilot Study of Implementing COVID-19 Mitigation Measures at an In-Person Conference"

To date, no study has audited ventilation and deployed auxiliary air purification during a conference. While the SSHI/SSH Safety Concept mentioned using "smart working ventilation," the specifics were not detailed [50]. At the time of the NAVSA conference, there was no uniform guidance for air quality mitigation. We consulted the emerging literature on SARS-CoV-2, ventilation, CO2 ppm, ACH, and purification [CDC 2021;Biro et al 2012; Batterman 2017; Di Gilio 2021, Gouge, 2023; McNeill 20220. We developed a system using CO2 sensors, Raspberry Pi computers, Corsi-Rosenthal boxes, and student volunteers (to count occupancy). If ACH fell below 5, a relay would trigger the Corsi-Rosenthal box to run for the duration of a panel session and provide estimated compensatory ACH equivalence. Unfortunately, the use of Google Sheets to centralize ACH calculations from each unit (10) relied on hotel Wi-Fi, which proved unstable. Consequently, the organizers decided to leave the computer units on for data collection but manually turned all Corsi-Rosenthal boxes on with wired bypass switch. While we were unable to measure the effect of risk reduction from purifiers, the intervention was welcomed by the majority (67%) of participants (Table 2). As a note, the CDC recently updated and expanded their ventilation guidelines (May 12, 2023), specifically recommending targeting 5 ACH in buildings and adding a section on the use of "DIY Air Cleaners" (e.g. Corsi-Rosenthal Boxes) [30]. Current experimental work on DIY purification continues to support their efficacy while specifying limitations based on build and configuration (Aldekheel, et al 2022;Cadnum et al 2015)

$$A_s = 6 \times 10^4 n G n_p / \left[ V (C_s - C_r) \right] \tag{B1}$$

Steady-state CO2 Air Change Rate

where $A_s$ is the steady-state air change rate,$n$ is the number of people occupying the space, $G_p$ is the average CO2 generation rate per person in units of Liters divided by min and number of people, $V$ is the volume of the room in units of cubic meters, $C_s$ and $C_r$ are the steady-state indoor CO2 concentration and CO2 concentration in replacement of outdoor air both in units of parts per million. The estimate of $G_p$ assumed 30-40 males (margin of overestimation for precautionary reasons) at 1.5 metabolic equivalents (METS) of energy expenditure, yielding 0.333 L/min/person) (Persily and Jong 2017).

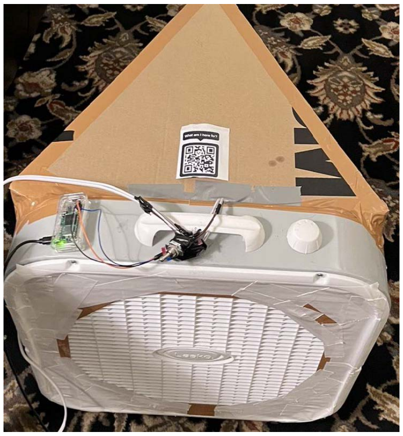

FIG. S1. Modified Corsi-Rosenthal Box: (i) one 25-inch Lasko Fan; (ii) two 3M MERV 13 Filters; (iii) one Raspberry Pi computer with Bluetooth and Wi-Fi; (iv) 5-volt relay triggered by Raspberry Pi Python program monitoring CO2 data from Aranet CO2 Monitor via Bluetooth (not pictured) (v) back up hardwired switch

| | | | | | | |
|---|---|---|---|---|---|---|
| Room | Lafayette | | | | | |
| Formula Assumptions: | | | | | | |
| Corsi box CADR (CFM) | 350 | CO2 generation rate (L/m) | 0.333 | | | |
| No. People | 10 | Outdoor CO2 PPM | 450.667 | | | |
| Room area (sq ft) | 496 | Room volume (ft^3) | 4960 | | | |
| Room height (ft) | 10 | Room volume (m^3) | 140.4515591 | | | |
| Corsi boxes used | 1 | ACH produced per box | 3.87 | | | |
| Session | Occupancy | Date Time | CO2 (ppm) | Mac Address | ACH without box | ACH with box(es) on |
| 3B: Radical Visions | 10 | 09-29-2022 1:38 PM | 2034 | 60:C0:BF:48:D0:19 | 1.8868 | 5.7577 |
| 3B: Radical Visions | 10 | 09-29-2022 1:43 PM | 2011 | 60:C0:BF:48:D0:19 | 1.9146 | 5.7855 |
| 3B: Radical Visions | 10 | 09-29-2022 1:48 PM | 1929 | 60:C0:BF:48:D0:19 | 2.0208 | 5.8917 |
| 3B: Radical Visions | 10 | 09-29-2022 1:53 PM | 1952 | 60:C0:BF:48:D0:19 | 1.9898 | 5.8608 |
| 3B: Radical Visions | 10 | 09-29-2022 1:58 PM | 2041 | 60:C0:BF:48:D0:19 | 1.8785 | 5.7494 |
| 3B: Radical Visions | 10 | 09-29-2022 2:03 PM | 2076 | 60:C0:BF:48:D0:19 | 1.8380 | 5.7090 |
| 3B: Radical Visions | 10 | 09-29-2022 2:08 PM | 2077 | 60:C0:BF:48:D0:19 | 1.8369 | 5.7078 |
| 3B: Radical Visions | 10 | 09-29-2022 2:13 PM | 2108 | 60:C0:BF:48:D0:19 | 1.8025 | 5.6735 |
| 3B: Radical Visions | 10 | 09-29-2022 2:18 PM | 1982 | 60:C0:BF:48:D0:19 | 1.9508 | 5.8218 |
| 3B: Radical Visions | 10 | 09-29-2022 2:23 PM | 1973 | 60:C0:BF:48:D0:19 | 1.9624 | 5.8333 |
| 3B: Radical Visions | 10 | 09-29-2022 2:28 PM | 1988 | 60:C0:BF:48:D0:19 | 1.9432 | 5.8142 |

FIG. S2. Sample Google Sheet Data from an operational automatic trigger and data log. Upon initial reading, ACH was 1.88 (<5), which triggered the Python script to turn on Raspberry Pi GPIO pins connected to the relay (set to normally off [NO]) and completed the circuit to turn on the fan. In the sample room and panel above, the purification added the equivalent of 3.87 ACH, putting the room above desired 5 ACH.
