## Appendix C: Survey Instruments for "Can a Conference be Epidemiologically Conscious? A Pilot Study of Implementing COVID-19 Mitigation Measures at an In-Person Conference"

Survey Questions

Below are survey questions for each type of survey: (i) informed consent + pre-survey; (ii) daily antigen check-in; (iii) post-conference de-brief; (iv) post-conference antigen check-in; (v) informed-consent + positivity confirmation. For brevity, only one daily antigen check-in (of 4) and one post-conference (of 5) antigen daily check-in have been included. Please see Supplemental Figure S1 for study flow diagram.

### Informed Consent -> Presurvey

---

Start of Block: Informed Consent

Q31

**Informed Consent**

**Title**

Conference attendance and COVID-19 behaviors

**Names of Principal Investigator**

Dr. Lorenzo Servitje, Associate Professor, Lehigh University

**Brief Description**

A series of online surveys will be administered using Qualtrics survey software to determine if specific COVID mitigation practices may modulate perceived risk among people who are attending the in-person conference.

**Procedure**

Participants who opt-in into the survey will receive personalized links to 4 different surveys.
Generally, they will ask questions about behaviors, perception, use of masks, and use and results of antigen tests. Participants will be asked to use rapid tests and report results but can decline to answer if they so choose; participants can choose not to test.

The Pre-Conference Survey links will be sent 5 days before the conference (9/23) and will close on day 1 of the conference (9/28). Daily email reminders will be sent to those who opt in at 9 AM each day. Surveys will include questions related to health and risk reduction behaviors such as masking, knowledge surrounding COVID-19, travel behavior and demographic questions; questions about self-assessed health status, vaccination and COVID-19 diagnosis.

Daily-Check-ins will be sent out every day of the conferences (9/28-10/2). Based on opt-in information, participants will receive links beginning on their first morning at the conference. We ask that participants test first thing in the morning. Surveys will be sent at 7:30 AM; we suggest participants schedule 15-20 minutes for antigen tests before sessions (first event is 8:30), while the survey itself should take no more than 3-5 minutes. Noon and evening reminders (8 PM) will be sent. Surveys will include questions surrounding rapid test completion, results, signs and symptoms, and activities attended the day previously. If participants have not completed a pre-survey, demographic and vaccination history questions from the pre-conference survey will be included in subsequent surveys.

Post Conference Debrief link will be sent out on the last day of the conference (10/2) and close 3 days afterwards (10/5). Daily reminders will be sent to those who opt in at 9 AM each day. Questions will include perceptions of risk during the conference; open-ended questions related to COVID-19 mitigation strategies and the effect on conference experience; questions about the use of masks and tests; questions related to knowledge of COVID-19. If participants have not completed a pre-survey, demographic and vaccination history questions from pre-conference survey will be included.

Post Conference Antigen Survey reminders will be sent one day before a participant's day of departure to confirm they have picked up 5 free antigen tests. Surveys will be sent out one day after participants departure at 8:30 AM with noon and evening reminders (8 PM). Questions will include completion and results of rapid antigen tests, signs and symptoms, along with known exposures during or after the conference. If participants have not completed a pre-survey, demographic and vaccination history questions from Pre-conference survey will be included.

**Potential Risks**
There may be some emotions that surface regarding negative experiences with COVID-19 or with COVID-19 mitigation practices. Some of the survey questions may trigger emotional or mental distress. There is a potential risk of identifying participants by the email address as they enter the survey,

**Protection against Risks**
At any point in the survey, participants may select "prefer not to answer," and/or simply exit the survey if desired.

To mitigate this risk of identification, email addresses will be collected through the Qualtrics participation survey that assigns a random number ID to each email address, storing the pairings in a Qualtrics directory to which only the PI will have access and which is protected on the Qualtrics website via password protection. Once all of the surveys have been completed, the responses will be de-identified prior to analysis, and there will be no way to determine whose responses are being analyzed.

**Potential Benefits**
Questions surrounding mask use, antigen testing, and risk reduction behaviors have become fairly standard in daily living and working. There is currently minimal information about how and why individuals behave the way they do in response to COVID-19 mitigation procedures at in-person conferences. By analyzing the responses of individuals at the conference, the benefits and challenges concerning mitigation efforts can be identified, enabling strategies to be designed to help improve future in-person conferences by making them more safe and comfortable.

**Compensation for Participation**
Participants will be given instructions regarding payment at the beginning of each survey.
Presurvey payments will be paid with 24 hours via Amazon gift sent digitally via email (participants will be given to links at the end of surveys linking to an external survey to prevent linking responses to their payment email).
Presurvey: $10
Daily Check-In: $5 per day. $10 bonus completing daily check in days.
Post Survey Debrief: $10
Post Conference Antigen Survey: $5 per day; $10 bonus completing all days.
Bonus completion for all surveys (1-4): $10
Interview: $20 (separate consent required)

*Delivery of gift cards*
Delivery of gift cards for Presurvey will be delivered within 24 hours of email being entered into post-survey google form. On-site or remote interview payments will be paid upon completion of interview by transferring the

digital amazon card to the email of participant's choice. Daily Check-in, Post-Survey Debrief, Post Conference-Antigen tests, and bonus gift cards will be sent no later than 3 weeks after the completion of data collection (10/31).

**Alternatives to Participation**
This is a completely voluntary survey, and those who feel uncomfortable at any time do not need to take it or can stop at any point. Provision of antigen tests, masks, COVID-19 information during the conference is free of charge and participation in the surveys is in no way required.

**Information withheld**
There will be no information withheld from individuals participating in any part of this study.

**Confidentiality**
All information will remain confidential in the process of collecting responses during the survey period. Email addresses will be collected using an interest-to-participate survey Qualtrics survey. A random number ID will then be assigned to each email address, and the pairings will be stored in a Qualtrics directory to which only the PI will have access. This allows Qualtrics to generate personalized links which will be sent to each participant ID to enter the first survey and those that follow. The personalized URLs linked to random IDs removes the need to enter any identifying information during the surveys themselves, but also allows previous responses to questions (such as demographics) to be carried over, eliminating the need to for participants to answer them again.

The Qualtrics directory is password protected and only accessible to the PI. Once all of the surveys have been completed, the responses will be de-identified prior to analysis, and there will be no way to determine whose responses are being analyzed. While it is very unlikely, and every safeguard has been placed to prevent this, a data breach is possible.

---

Q36 Do you agree and consent to participate in this study?

- ○ Yes (2)
- ○ No (1)

End of Block: Informed Consent

---

Start of Block: Travel Plans

Q29 Thank you for participating in the pre-conference survey. This survey should take you no more than 5-10 minutes.

Please make sure to click the link at the end of the survey to go to an external Google form where you can enter the email address where you would like your Amazon gift card delivered.

---

Q1 How far will you travel or did you travel to attend this conference?

- ○ Less than 25 miles (1)
- ○ 26-100 miles (2)
- ○ 101-250 miles (3)
- ○ 251-500 miles (4)
- ○ More than 500 miles (5)
- ○ Prefer not to answer (6)

---

Q2 By what means will you or did you travel? (Select all that apply)

- ☐ Alone in a private vehicle (1)
- ☐ With 1-2 other people in a private vehicle (2)
- ☐ Bus (3)
- ☐ Train (4)
- ☐ Airplane (5)
- ☐ Taxi/Uber/Lyft (6)
- ☐ Prefer not to answer (8)
- ☐ Other (please specify) (7) ________________________________________________

---

Q3 In the week prior to traveling to the conference, which of the following have you or did you engage in? (Select all that apply)

- ☐ Going on a walk (1)
- ☐ Attended an indoor event with people I do not live with (2)
- ☐ Attended a movie/concert/play (3)
- ☐ Grocery store shopping (4)
- ☐ Eaten inside at a restaurant (5)
- ☐ Traveled on public transportation (6)
- ☐ Filled up a car with gas at a gas station (7)
- ☐ None of these (9)
- ☐ Prefer not to answer (8)

Q4 In the week prior to the conference, which COVID prevention behaviors did you or will you generally practice? (Select all that apply)

- ☐ Wearing a well-fitting mask in all outdoor public spaces (1)
- ☐ Wearing a well-fitting mask in all indoor public spaces (2)
- ☐ Avoid large gatherings of people (3)
- ☐ Test for COVID if symptomatic (4)
- ☐ Test for COVID if known exposure (5)
- ☐ Isolate if COVID positive until testing negative (6)
- ☐ Isolate if COVID positive for 5 days (7)
- ☐ Notify close contacts of potential exposure to COVID (8)
- ☐ Disinfecting packages and/or mail (9)
- ☐ None of these (11)
- ☐ Prefer not to answer (12)
- ☐ Other (please specify) (10) ________________________________________________

Q5 What COVID precautions will you or did you take on your trip to the conference? (Select all that apply)

- ☐ Wear a well-fitting mask whenever inside/indoors (1)
- ☐ Wear a well-fitting mask whenever outdoors (2)
- ☐ Test prior to attending the conference (3)
- ☐ Test during the conference (4)
- ☐ Test prior to returning home (5)
- ☐ Test 3-5 days after returning home (6)
- ☐ None of these (7)
- ☐ Prefer not to answer (8)

End of Block: Travel Plans

---

Start of Block: Health, Household, Risk

Q6 How would you rate your overall health?

- ○ Excellent (1)
- ○ Good (2)
- ○ Fair (3)
- ○ Poor (4)
- ○ Other (please specify) (5) ________________________________________________
- ○ Prefer not to answer (6)

---

Q7 Which, if any, of the following categories apply to you?

- ☐ Over the age of 65 (1)
- ☐ Immunocompromised (2)
- ☐ Afflicted with underlying health conditions (e.g., cardiovascular disease, diabetes, etc.) that make them more vulnerable to COVID-19 (3)
- ☐ Pregnant or recently pregnant (4)
- ☐ I do not fall into any of these categories (8)
- ☐ Prefer not to answer (7)

---

Q33 Which, if any, of the following categories apply to anyone else in your household?

- ☐ Over the age of 65 (1)
- ☐ Immunocompromised (2)
- ☐ Afflicted with underlying health conditions (e.g., cardiovascular disease, diabetes, etc.) that make them more vulnerable to COVID-19 (3)
- ☐ Pregnant or recently pregnant (4)
- ☐ No one else falls into any of these categories (8)
- ☐ Prefer not to answer (7)

---

Page Break

Q9 At this point, how concerned are you about the risk COVID-19 poses to you or anyone in your household?

○ Extremely concerned (49)

○ Somewhat concerned (50)

○ Neither concerned nor unconcerned (51)

○ Somewhat unconcerned (52)

○ Extremely unconcerned (53)

○ Prefer not to answer (54)

---

Q11 How would you rate the risk of attending this conference in general?

○ Extremely risky (38)

○ Somewhat risky (39)

○ Neither risky nor safe (40)

○ Somewhat safe (41)

○ Extremely safe (42)

○ Prefer not to answer (43)

---

Q12 Compared to your activities of daily life and work, how would you characterize the risk of attending this conference?

○ Conference is less risky (1)

○ Conference is the same level of risk (2)

○ Conference is more risky (3)

○ Prefer not to answer (4)

**End of Block: Health, Household, Risk**

---

**Start of Block: COVID Knowledge/Belief**

Q37 Thank you. The next five questions ask you to rate statements about COVID-19 as "True" or "False." Please indicate, to the best of your knowledge, your belief about each statement.

---

Q13 SARS-CoV-2 is a virus that can be transmitted through the air.

- ○ Definitely true (4)
- ○ Probably true (3)
- ○ Probably false (2)
- ○ Definitely false (1)
- ○ Prefer not to answer (5)

---

Q14 It is not possible to transmit the virus 5 days after a positive test.

- ○ Definitely true (5)
- ○ Probably true (6)
- ○ Probably false (8)
- ○ Definitely false (9)
- ○ Prefer not to answer (10)

---

Q15 It is not possible to be re-infected with the virus.

- ○ Definitely true (5)
- ○ Probably true (6)
- ○ Probably false (8)
- ○ Definitely false (9)
- ○ Prefer not to answer (10)

Q16 Being up-to-date on COVID vaccinations protects against severe disease.

- Definitely true (5)
- Probably true (6)
- Probably false (8)
- Definitely false (9)
- Prefer not to answer (10)

Q19 If you are up-to-date on vaccinations, there are no additional benefits from wearing a mask.

- Definitely true (5)
- Probably true (6)
- Probably false (8)
- Definitely false (9)
- Prefer not to answer (10)

End of Block: COVID Knowledge/Belief

Start of Block: COVID Vaccination/History

Q18 Have you been vaccinated for COVID-19?

- Yes (1)
- No (2)
- Prefer not to answer (3)

*Display This Question:*

*If Have you been vaccinated for COVID-19? = Yes*

Q20 Which statement describes your current vaccine status? (Select all that apply)

- ☐ I have received 1 dose of the Johnson and Johnson vaccine only (1)
- ☐ I have received 1 dose of the Johnson and Johnson vaccine and one Booster (2)
- ☐ I have received 1 dose of the Pfizer/Moderna vaccine (3)
- ☐ I have received 2 doses of the Pfizer/Moderna vaccine (4)
- ☐ I have received 3 doses of the Pfizer/Moderna vaccine (5)
- ☐ I have received 4 doses of the Pfizer/Moderna vaccine (original, not bivalent) (6)
- ☐ I have received the Pfizer/Moderna Bivalent Booster (9)
- ☐ Other (please specify) (7) ________________________________________________
- ☐ Prefer not to answer (10)

---

*Display This Question:*

*If Have you been vaccinated for COVID-19? = Yes*

Q21 What was the date of your most recent vaccination/booster?

- ○ Within the last 2 weeks (1)
- ○ More than 2 weeks but less than 2 months (2)
- ○ 2-6 months ago (3)
- ○ More than 6 months ago (4)
- ○ I have not received any booster vaccinations (5)
- ○ Other (please specify) (6) ________________________________________________

---

Q22 Have you ever tested positive for Covid-19?

- ◯ Yes, I tested positive on a home test (rapid test/antigen test: Flowflex, Binaxtest, or similar; please approximate provide the date) (2) ________________________________________________

- ◯ Yes, I tested positive on a test given to me by a doctor or health care professional (PCR test) on (please approximate provide the date) (3) ________________________________________________

- ◯ Yes, other (1) ________________________________________________

- ◯ No, I have not tested positive (4)

- ◯ Prefer not to answer (5)

---

Q23 Regardless of any testing you've had done, do you believe you've have COVID?

- ◯ Yes (please provide the month and year) (1) ________________________________________________

- ◯ No (2)

- ◯ Unsure (3)

- ◯ Prefer not to answer (4)

End of Block: COVID Vaccination/History

---

Start of Block: Demographic Questions

Q24 Please type your age below in numbers (i.e., "48"). If you do not want to reply, please type "prefer not to answer"

________________________________________________________________

---

Q25 With which gender do you identify?

- ○ Man (1)
- ○ Woman (2)
- ○ Non-binary / third gender (3)
- ○ Prefer to self-describe (Please specify) (4) ________________________________________________
- ○ Prefer not to answer (5)

---

Q26 What is your race / ethnicity? (Please select all that apply)

- ▢ Asian / Pacific Islander (1)
- ▢ Black / African American (2)
- ▢ Hispanic / Latino (3)
- ▢ Native American / American Indian (4)
- ▢ White / Caucasian (5)
- ▢ Prefer to self-describe (Please specify) (6) ________________________________________________
- ▢ Prefer not to answer (7)

---

Q27 What is your highest level of education?

◯ High School Diploma (1)

◯ Bachelor's Degree (2)

◯ Master's Degree (3)

◯ Doctorate (4)

◯ Other (Please specify) (5) ________________________________________________

◯ Prefer not to answer (6)

---

Q28 What is your professional status?

◯ Tenure track, Assistant (1)

◯ Tenure track, Associate (2)

◯ Tenure track, Professor (3)

◯ Graduate Student (4)

◯ Non-tenure track (5)

◯ Prefer to self-describe (Please specify) (6) ________________________________________________

◯ Prefer not to answer (7)

**End of Block: Demographic Questions**

---

**Start of Block: Block 6**

Right Answers Before concluding your survey, please consult the answers to the COVID knowledge questions from the earlier part of the survey:

1. "SARS-CoV-2 is a virus that can be transmitted through the air." You answered: ${Q13/ChoiceGroup/SelectedChoices}

*SARS-CoV-2, or the virus that causes COVID-19,* ***can indeed be transmitted through the air.*** *The virus is easily spread by aerosols exhaled by an infected individual. Aerosols can travel through the air like cigarette smoke.*

2. "It is not possible to transmit the virus 5 days after a positive test." You

answered: ${Q14/ChoiceGroup/SelectedChoices}

***It is possible to transmit the virus 5 days after a positive rapid antigen test.*** *There has been a lot of misinformation about the issue of how long a person is able to transmit COVID-19 after a positive test. Many public health experts agree that 5 days is not enough time for a person to safely return to their normal activities, as they may still be able to infect others.*

3. "It is not possible to be re-infected with the virus." You answered: ${Q15/ChoiceGroup/SelectedChoices}

*With extant variants, an* ***infection with COVID-19 does not create long-term immunity, and it is possible to be re-infected.***

4. "Being up-to-date on COVID vaccinations protects against severe disease." You answered: ${Q16/ChoiceGroup/SelectedChoices}

*The data show that* ***people who are up-to-date on vaccinations are much less likely to experience severe disease, hospitalization, or death.***

5. "If you are up-to-date on vaccination, there are no additional benefits from mask." You answered: ${Q19/ChoiceGroup/SelectedChoices}

*Though* ***vaccination is important, it does not offer perfect protection against COVID-19. Wearing a well-fitting mask can help vaccinated people reduce their chances of infection.***

End of Block: Block 6

---

### Daily Check-in Survey 9/30

---

Start of Block: Default Question Block

Q8 Good morning! Thank you for participating in our daily check-in. You will be asked about activities from Thursday, any signs or symptoms, and the results of your antigen test. If you have not completed your rapid test, it might be a good time to so now (results take 15 minutes).

Please make sure to click the link at the end of the survey to go to an external Google form where you can enter the email address where you would like your Amazon gift card delivered.

The survey that follows only takes 3-5 minutes.

Q7 If you attended any of the following events on Thursday 9/29, please select which of the following apply.

| | Wore a mask indoors at all times when around other people (1) | Wore a mask indoors sometimes when around other people (2) | Used hand sanitizer or washed hands before eating or after touching shared objects (3) | Ate and/or socialized outdoors (4) | None of these (5) | Prefer not answer (6) |
|---|---|---|---|---|---|---|
| Breakfast (1) | ▢ | ▢ | ▢ | ▢ | ▢ | ▢ |
| 1A: Not Just Victorianists (2) | ▢ | ▢ | ▢ | ▢ | ▢ | ▢ |
| 1B: Teaching More than Just Music or Literature (34) | ▢ | ▢ | ▢ | ▢ | ▢ | ▢ |
| 1C: (Un)Just Networks (3) | ▢ | ▢ | ▢ | ▢ | ▢ | ▢ |
| 1D: Just in Time: The Victorian Present & Non-Linear Temporalities (4) | ▢ | ▢ | ▢ | ▢ | ▢ | ▢ |
| 1E: Doing Justice to Teaching East Asia in Victorian Studies (5) | ▢ | ▢ | ▢ | ▢ | ▢ | ▢ |
| 1F: Storied Afterlives (75) | ▢ | ▢ | ▢ | ▢ | ▢ | ▢ |
| 2A: Body Parts (6) | ▢ | ▢ | ▢ | ▢ | ▢ | ▢ |
| 2B: “British” Flora: The Politics of Plants in Victorian Art & Science (7) | ▢ | ▢ | ▢ | ▢ | ▢ | ▢ |
| 2C: Not Just Care (8) | ▢ | ▢ | ▢ | ▢ | ▢ | ▢ |

| | | | | | | |
|---|---|---|---|---|---|---|
| 2D: Monsters, Mimics, Martyrs, & Jews: The Radical Imaginaries of British India (9) | ☐ | ☐ | ☐ | ☐ | ☐ | ☐ |
| 2E: Newspapers and Empire (51) | ☐ | ☐ | ☐ | ☐ | ☐ | ☐ |
| 2F: Staging Justice (52) | ☐ | ☐ | ☐ | ☐ | ☐ | ☐ |
| 2G: The Normal and the Objectionable (53) | ☐ | ☐ | ☐ | ☐ | ☐ | ☐ |
| Complimentary Conference Lunch (62) | ☐ | ☐ | ☐ | ☐ | ☐ | ☐ |
| Teaching Universities Caucus (76) | ☐ | ☐ | ☐ | ☐ | ☐ | ☐ |
| Data Caucus (77) | ☐ | ☐ | ☐ | ☐ | ☐ | ☐ |
| Victorian Poetry Caucus (78) | ☐ | ☐ | ☐ | ☐ | ☐ | ☐ |
| Victorian Art History Caucus (79) | ☐ | ☐ | ☐ | ☐ | ☐ | ☐ |
| Nineteenth-Century Theatre Caucus (80) | ☐ | ☐ | ☐ | ☐ | ☐ | ☐ |
| 3A: Re-presenting Victorian Studies (55) | ☐ | ☐ | ☐ | ☐ | ☐ | ☐ |
| 3B: Radical Visions (56) | ☐ | ☐ | ☐ | ☐ | ☐ | ☐ |
| 3C: Dirty Humanities (57) | ☐ | ☐ | ☐ | ☐ | ☐ | ☐ |
| 3D: (Rethinking) Marriage Plots (58) | ☐ | ☐ | ☐ | ☐ | ☐ | ☐ |

| | | | | | | |
|---|---|---|---|---|---|---|
| 3E: Just Data: Working with Datasets in Victorian Studies (59) | ☐ | ☐ | ☐ | ☐ | ☐ | ☐ |
| 3F: Aesthetics of Justice (60) | ☐ | ☐ | ☐ | ☐ | ☐ | ☐ |
| 4A: Religious Justice (61) | ☐ | ☐ | ☐ | ☐ | ☐ | ☐ |
| 4B: Not Just Art for Art's Sake (63) | ☐ | ☐ | ☐ | ☐ | ☐ | ☐ |
| 4C: Pioneers, Touch Tours, and Totems (64) | ☐ | ☐ | ☐ | ☐ | ☐ | ☐ |
| 4D: Just in Time (65) | ☐ | ☐ | ☐ | ☐ | ☐ | ☐ |
| 4E: Information and Sensory Overload (66) | ☐ | ☐ | ☐ | ☐ | ☐ | ☐ |
| 4F: Crime and Punishment (67) | ☐ | ☐ | ☐ | ☐ | ☐ | ☐ |
| 4G: Seminar 1: Care Ethics and Victorian Fiction (68) | ☐ | ☐ | ☐ | ☐ | ☐ | ☐ |
| Plenary Address (69) | ☐ | ☐ | ☐ | ☐ | ☐ | ☐ |
| Opening Reception (70) | ☐ | ☐ | ☐ | ☐ | ☐ | ☐ |

Q4 Did you experience any of the following symptoms yesterday or this morning? (Please select all that apply)

- ☐ Fever (1)
- ☐ Headache (2)
- ☐ Sore throat (3)
- ☐ Shortness of breath (4)
- ☐ Cough (5)
- ☐ Muscle aches (6)
- ☐ Extreme fatigue (7)
- ☐ Nausea / vomiting (8)
- ☐ Loss of taste or smell (9)
- ☐ Other (Please specify) (10) ________________________________________________
- ☐ I did not have any of these symptoms today (11)
- ☐ Prefer not to answer (13)

---

`

Q5 Did you take a rapid COVID test today (i.e., Flowflex, Binax test, or similar)?

- ○ Yes (1)
- ○ No (2)
- ○ Prefer not to answer (3)

---

*Display This Question:*

*If Did you take a rapid COVID test today (i.e., Flowflex, Binax test, or similar)? = Yes*

Q6 What was the result?

◯ Negative (one line) (1)

◯ Positive (two lines, including a reading of two faint lines) (2)

◯ I don't know (3)

◯ Prefer not to answer (4)

---

*Display This Question:*

*If What was the result? = Positive (two lines, including a reading of two faint lines)*

Q9 We are sorry to hear you tested positive. We hope you contact your care provider and recover quickly. Please contact any possible contacts you may have exposed.

If you require assistance, please text the conference organizers (833) 676-3967 or send them an email and also consult the conference's "Getting Help" page.

For more information, please see NAVSA 2022's Covid Policies and Procedures.

We hope you feel better soon. Please follow the next link to complete your survey and enter your email for your amazon gift card.

End of Block: Default Question Block

---

### Final Post Conference Debrief

---

Start of Block: Travel Plans

Q46 Based on the information from the opt-in initial survey, you should be receiving this the day of or after your last day at the conference. We hope you had a good conference and safe travels home.

In this survey, you will be asked about the following: risk perceptions and behaviors during travel home and during the conference; open-ended questions about the COVID-19 mitigation strategies and conference experience; questions about the use of masks and tests; and questions related to knowledge of COVID-19.

This survey should take you no more than 10-15 minutes.

Please make sure to click the link at the end of the survey to go to an external Google form where you can enter the email address where you would like your Amazon gift card delivered.

Q1 What precautions did you take on your return from the conference? Please select all that apply.

- [ ] Wear a well-fitting mask whenever I am inside/indoors. (1)
- [ ] Wear a well-fitting mask whenever I am outdoors. (2)
- [ ] Test prior to attending the conference. (3)
- [ ] Testing prior to returning home (4)
- [ ] Testing upon return home (5)
- [ ] Other (6) ________________________________________________
- [ ] Prefer not to answer (8)

Q2 By what means did you travel? (Select all that apply)

- [ ] Alone in a private vehicle (1)
- [ ] With 1-2 other people in a private vehicle (2)
- [ ] Bus (3)
- [ ] Train (4)
- [ ] Airplane (5)
- [ ] Taxi/Uber/Lyft (6)
- [ ] Other (please specify) (7) ________________________________________________
- [ ] Prefer not to answer (8)

Q4 In the last week, which COVID-19 prevention behaviors have you generally practiced? (Select all that apply)

- ▢ Wearing a well-fitting mask in all outdoor public spaces (1)
- ▢ Wearing a well-fitting mask in all indoor public spaces (2)
- ▢ Avoid large gatherings of people (3)
- ▢ Test for COVID if symptomatic (4)
- ▢ Test for COVID if known exposure (5)
- ▢ Isolate if COVID positive until testing negative (6)
- ▢ Isolate if COVID positive for 5 days (7)
- ▢ Notify close contacts of potential exposure to COVID (8)
- ▢ Disinfecting packages and/or mail (9)
- ▢ Other (please specify) (10) ________________________________________________

End of Block: Travel Plans

---

Start of Block: Covid Knowledge/Belief

Q35 Please answer the following questions to the best of your knowledge.

Q13 SARS-CoV-2 is a virus that can be transmitted through the air.

- Definitely true (13)
- Probably true (14)
- Probably false (16)
- Definitely false (17)
- Prefer not to answer (18)

---

Q14 It is not possible to transmit the virus 5 days after a positive test.

- Definitely true (20)
- Probably true (21)
- Probably false (23)
- Definitely false (24)
- Prefer not to answer (25)

---

Q15 It is not possible to be re-infected with the virus.

- Definitely true (9)
- Probably true (10)
- Probably false (11)
- Definitely false (12)
- Prefer not to answer (14)

---

Q16 Being up-to-date on COVID vaccinations protects against severe disease.

- Definitely true (9)
- Probably true (10)
- Probably false (11)
- Definitely false (12)
- Prefer not to answer (14)

---

Q19 If you are up-to-date on vaccinations, there are no additional benefits from wearing a mask.

- Definitely true (17)
- Probably true (18)
- Probably false (19)
- Definitely false (20)
- Prefer not to answer (22)

End of Block: Covid Knowledge/Belief

---

Start of Block: Knowledge of Mitigation Measures

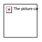

Q29 Which of the following COVID-19 mitigation measures did NAVSA 2022 employ (check all that apply)?

- ☐ Provide N95 masks free of charge before the conference (1)
- ☐ Require vaccination (2)
- ☐ Host a guest lecture on water infrastructure by John Snow (3)
- ☐ Provide estimated transmission and positivity rates of the conference population to attendees (4)
- ☐ Provide free rapid testing during the conference (5)
- ☐ Requested random rapid test screening (6)
- ☐ Upper Air UVC Lamp fixtures were installed in all conference rooms (7)
- ☐ Provide ventilation audits and information to participants (8)
- ☐ Provide free N95 masks during the conference (9)
- ☐ Provide local community transmission data (10)
- ☐ Send out Push notifications to "Mask Up" (12)
- ☐ I don't recall (11)

End of Block: Knowledge of Mitigation Measures

---

Start of Block: Health During Conference

Q4 Did you experience any of the following symptoms today? (Please select all that apply)

- ▢ Fever (1)
- ▢ Headache (2)
- ▢ Sore throat (3)
- ▢ Shortness of breath (4)
- ▢ Cough (5)
- ▢ Muscle aches (6)
- ▢ Extreme fatigue (7)
- ▢ Nausea / vomiting (8)
- ▢ Loss of taste or smell (9)
- ▢ Prefer not to answer (11)
- ▢ Other (Please specify) (10) ________________________________________________

---

Q30 Did you test positive for COVID-19 during or after the conference

- ○ Yes (2)
- ○ No (1)
- ○ Prefer not to answer (3)

---

*Display This Question:*

*If Did you test positive for COVID-19 during or after the conference = Yes*

Q47 When did you test positive? (please leave blank if you prefer not to answer)

________________________________________________________________

---

*Display This Question:*

*If Did you test positive for COVID-19 during or after the conference = Yes*

Q45 By what means did you confirm results?

- ◯ Rapid antigen test alone (1)
- ◯ Rapid antigen test and symptoms (2)
- ◯ PCR (3)
- ◯ Prefer not to answer (4)

**End of Block: Health During Conference**

---

**Start of Block: Risk Perception**

Q9 At this point, how concerned are you about the risk COVID-19 poses to you or anyone in your household?

- ◯ Extremely concerned (49)
- ◯ Somewhat concerned (50)
- ◯ Neither concerned nor unconcerned (51)
- ◯ Somewhat unconcerned (52)
- ◯ Extremely unconcerned (53)
- ◯ Prefer not to answer (54)

---

Q32 Please expand on your answer to the previous question.

________________________________________________________________

________________________________________________________________

________________________________________________________________

________________________________________________________________

________________________________________________________________

---

Q33 How would you rate the risk of attending this conference in general?

- ◯ Extremely risky (38)
- ◯ Somewhat risky (39)
- ◯ Neither risky nor safe (40)
- ◯ Somewhat safe (41)
- ◯ Extremely safe (42)
- ◯ Prefer not to answer (43)

---

Q34 Please expand on your answer to the previous question.

________________________________________________________________

________________________________________________________________

________________________________________________________________

________________________________________________________________

________________________________________________________________

---

Q36 Compared to your activities of daily life and work, how would you characterize the risk of attending this conference?

- ◯ Conference was less risky (1)
- ◯ Conference was the same level of risk (2)
- ◯ Conference was more risky (3)
- ◯ Prefer not to answer (4)

---

Q37 Please expand on your answer to the previous question.

________________________________________________________________

________________________________________________________________

________________________________________________________________

________________________________________________________________

________________________________________________________________

End of Block: Risk Perception

Start of Block: Block 8

Q38 Have you been to other in-person conferences since March 2020?

- Yes (1)
- No (2)
- Prefer not to answer (3)

*Display This Question:*

*If Have you been to other in-person conferences since March 2020? = Yes*

Q39 Were your behaviors and perceived risk similar or different? Please elaborate.

________________________________________________________________

________________________________________________________________

________________________________________________________________

________________________________________________________________

________________________________________________________________

Q40 Which individual provisions did you feel were helpful in reducing the risk of contracting an airborne infection like COVID-19?

- ▢ N95 masks (1)
- ▢ Access COVID-19 surveillance information about Bethlehem (2)
- ▢ Access to free rapid tests (3)
- ▢ Access to basic public health guidance information (4)
- ▢ Free PCR tests (6)
- ▢ Other (5) ________________________________________________

Page Break

Q41 Which structural provisions did you feel were helpful in reducing the risk of contracting an airborne infection like COVID-19?

- ▢ Encouraging and making freely accessible N95 masks to everyone (1)
- ▢ Encouraging and making freely accessible rapid tests everyone (2)
- ▢ Reducing attendee density via hybrid option (3)
- ▢ Air ventilation and measurement (4)
- ▢ Provision of COVID relief funds/hybrid presentation option for those who test positive before or during the conference (5)
- ▢ Upper Air UV Lamp fixtures (7)
- ▢ Other (6) ________________________________________________
- ▢ Prefer not to answer (8)

---

Q42 Did any COVID mitigations seem unnecessary, intrusive, obtrusive, or counterproductive? If so, how?

________________________________________________________________

________________________________________________________________

________________________________________________________________

________________________________________________________________

________________________________________________________________

---

Q43 How did you perceive other people's behavior around COVID precautions during the conference? Please feel free to share any general impressions, but please do not identify anybody individually or by name.

________________________________________________________________

________________________________________________________________

________________________________________________________________

________________________________________________________________

________________________________________________________________

Q44 Is there anything you would like to add about your experience at the conference? If so please share here.

________________________________________________________________

________________________________________________________________

________________________________________________________________

________________________________________________________________

________________________________________________________________

End of Block: Block 8

### Informed Consent -> Positivity

Start of Block: Welcome

Q44 Thank you for your interest in the NAVSA Study Positivity Confirmation Survey.

To be eligible, you must have been **1) physically present at the conference** *and* **2) have tested positive for COVID-19 either during the conference or the week following.**

In this survey, you will be asked to confirm the above, along with methods and dates of testing and possible exposures.

If you have previously completed surveys for the NAVSA Study, this should take you no more than 2-3 minutes to complete.

If you have not previously completed any surveys for the NAVSA Study, you will be asked another set of questions related to pre-conference and post-conference behaviors and risk perceptions questions, along with basic health and demographic data. This should take between 7-10 minutes to complete.

Please make sure to click the link at the end of the survey to go to an external Google form where you can enter the email address where you would like your Amazon gift card delivered.

End of Block: Welcome

Start of Block: Eligibility

Q64 Eligibility Questions:

Q1 Did you participate in NAVSA 2022 in-person?

- Yes (1)
- No (2)

Q2 Did you test positive for SARS-CoV2 during the conference (9/28-10/2) or during the week following the conference (10/3-10/10)?

- Yes (1)
- No (2)

Page Break

End of Block: Eligibility

Start of Block: Positivity

Q41 How did you test positive for SARS-CoV2?

- ◯ Rapid Antigen Test (Binax, Flowflex, or similar)  (1)
- ◯ PCR  (2)
- ◯ Prefer not to answer  (4)

Q40 What date did test positive for SARS-CoV2?

________________________________________________________________

Did you have  Did you have any known exposures you were are aware of? If so, what where they? (If you prefer not to answer, please leave blank)

________________________________________________________________

End of Block: Positivity

Start of Block: Health, Household, Risk

Q1 How far did  you travel to attend this conference?

- ◯ Less than 25 miles  (1)
- ◯ 26-100 miles  (2)
- ◯ 101-250 miles  (3)
- ◯ 251-500 miles  (4)
- ◯ More than 500 miles  (5)
- ◯ Prefer not to answer  (6)

Q3 In the week prior to traveling to the conference, which of the following did you engage in? (Select all that apply)

- ☐ Going on a walk (1)
- ☐ Attended an indoor event with people I do not live with (2)
- ☐ Attended a movie/concert/play (3)
- ☐ Grocery store shopping (4)
- ☐ Eaten inside at a restaurant (5)
- ☐ Traveled on public transportation (6)
- ☐ Filled up a car with gas at a gas station (7)
- ☐ None of these (9)
- ☐ Prefer not to answer (8)

Q4 In the week prior to the conference, which COVID prevention behaviors did you generally practice? (Select all that apply)

- [ ] Wearing a well-fitting mask in all outdoor public spaces (1)
- [ ] Wearing a well-fitting mask in all indoor public spaces (2)
- [ ] Avoid large gatherings of people (3)
- [ ] Test for COVID if symptomatic (4)
- [ ] Test for COVID if known exposure (5)
- [ ] Isolate if COVID positive until testing negative (6)
- [ ] Isolate if COVID positive for 5 days (7)
- [ ] Notify close contacts of potential exposure to COVID (8)
- [ ] Disinfecting packages and/or mail (9)
- [ ] None of these (11)
- [ ] Prefer not to answer (12)
- [ ] Other (please specify) (10) ________________________________________________

Q5 What COVID precautions did you take during your trip to the conference? (Select all that apply)

- ☐ Wear a well-fitting mask whenever inside/indoors (1)
- ☐ Wear a well-fitting mask whenever outdoors (2)
- ☐ Test prior to attending the conference (3)
- ☐ Test during the conference (4)
- ☐ Test prior to returning home (5)
- ☐ Test 3-5 days after returning home (6)
- ☐ None of these (7)
- ☐ Prefer not to answer (8)

---

Page Break

Q6 How would you rate your overall health?

- ○ Excellent (1)
- ○ Good (2)
- ○ Fair (3)
- ○ Poor (4)
- ○ Other (please specify) (5) ________________________________________________
- ○ Prefer not to answer (6)

Q7 Which, if any, of the following categories apply to you?

- ▢ Over the age of 65 (1)
- ▢ Immunocompromised (2)
- ▢ Afflicted with underlying health conditions (e.g., cardiovascular disease, diabetes, etc.) that make them more vulnerable to COVID-19 (3)
- ▢ Pregnant or recently pregnant (4)
- ▢ I do not fall into any of these categories (8)
- ▢ Prefer not to answer (7)

Q33 Which, if any, of the following categories apply to anyone else in your household?

- ▢ Over the age of 65 (1)
- ▢ Immunocompromised (2)
- ▢ Afflicted with underlying health conditions (e.g., cardiovascular disease, diabetes, etc.) that make them more vulnerable to COVID-19 (3)
- ▢ Pregnant or recently pregnant (4)
- ▢ No one else falls into any of these categories (8)
- ▢ Prefer not to answer (7)

---

Page Break

End of Block: Health, Household, Risk

Start of Block: COVID Vaccination/History

Q18 Have you been vaccinated for COVID-19?

- Yes (1)
- No (2)
- Prefer not to answer (3)

*Display This Question:*

*If Have you been vaccinated for COVID-19? = Yes*

Q20 Which statement describes your current vaccine status? (Select all that apply)

- I have received 1 dose of the Johnson and Johnson vaccine only (1)
- I have received 1 dose of the Johnson and Johnson vaccine and one Booster (2)
- I have received 1 dose of the Pfizer/Moderna vaccine (3)
- I have received 2 doses of the Pfizer/Moderna vaccine (4)
- I have received 3 doses of the Pfizer/Moderna vaccine (5)
- I have received 4 doses of the Pfizer/Moderna vaccine (original, not bivalent) (6)
- I have received the Pfizer/Moderna Bivalent Booster (9)
- Other (please specify) (7) ________________________________________________
- Prefer not to answer (10)

*Display This Question:*

*If Have you been vaccinated for COVID-19? = Yes*

Q21 What was the date of your most recent vaccination/booster?

- ○ Within the last 2 weeks (1)
- ○ More than 2 weeks but less than 2 months (2)
- ○ 2-6 months ago (3)
- ○ More than 6 months ago (4)
- ○ I have not received any booster vaccinations (5)
- ○ Other (please specify) (6) ________________________________________________

---

Q22 Prior to this last positive test you reported above, had you ever tested positive for Covid-19?

- ○ Yes, I tested positive on a home test (rapid test/antigen test: Flowflex, Binaxtest, or similar; please approximate provide the date) (2) ________________________________________________
- ○ Yes, I tested positive on a test given to me by a doctor or health care professional (PCR test) on (please approximate provide the date) (3) ________________________________________________
- ○ Yes, other (1) ________________________________________________
- ○ No, I have not tested positive (4)
- ○ Prefer not to answer (5)

---

Q23 Regardless of any testing you've had done, do you believe you've have COVID?

- ○ Yes (please provide the month and year) (1) ________________________________________________
- ○ No (2)
- ○ Unsure (3)
- ○ Prefer not to answer (4)

End of Block: COVID Vaccination/History

---

Start of Block: Post Conf Qual

Q1 What precautions did you take on your return from the conference? Please select all that apply.

- ▢ Wear a well-fitting mask whenever I am inside/indoors. (1)
- ▢ Wear a well-fitting mask whenever I am outdoors. (2)
- ▢ Test prior to attending the conference. (3)
- ▢ Testing prior to returning home (4)
- ▢ Testing upon return home (5)
- ▢ Other (6) ________________________________________________
- ▢ Prefer not to answer (8)

---

Q4 In the week **during and after** the conference, which COVID-19 prevention behaviors did you generally practiced? (Select all that apply)

- ▢ Wearing a well-fitting mask in all outdoor public spaces (1)
- ▢ Wearing a well-fitting mask in all indoor public spaces (2)
- ▢ Avoid large gatherings of people (3)
- ▢ Test for COVID if symptomatic (4)
- ▢ Test for COVID if known exposure (5)
- ▢ Isolate if COVID positive until testing negative (6)
- ▢ Isolate if COVID positive for 5 days (7)
- ▢ Notify close contacts of potential exposure to COVID (8)
- ▢ Disinfecting packages and/or mail (9)
- ▢ Other (please specify) (10) ________________________________________________

Q9 At this point, how concerned are you about the risk COVID-19 poses to you or anyone in your household?

- ◯ Extremely concerned (49)
- ◯ Somewhat concerned (50)
- ◯ Neither concerned nor unconcerned (51)
- ◯ Somewhat unconcerned (52)
- ◯ Extremely unconcerned (53)
- ◯ Prefer not to answer (54)

Q32 Please expand on your answer to the previous question.

________________________________________________________________

________________________________________________________________

________________________________________________________________

________________________________________________________________

________________________________________________________________

Q33 How would you rate the risk of attending this conference in general?

- ◯ Extremely risky (38)
- ◯ Somewhat risky (39)
- ◯ Neither risky nor safe (40)
- ◯ Somewhat safe (41)
- ◯ Extremely safe (42)
- ◯ Prefer not to answer (43)

Q34 Please expand on your answer to the previous question.

________________________________________________________________

________________________________________________________________

________________________________________________________________

________________________________________________________________

________________________________________________________________

---

Q36 Compared to your activities of daily life and work, how would you characterize the risk of attending this conference?

◯ Conference was less risky (1)

◯ Conference was the same level of risk (2)

◯ Conference was more risky (3)

◯ Prefer not to answer (4)

---

Q37 Please expand on your answer to the previous question.

________________________________________________________________

________________________________________________________________

________________________________________________________________

________________________________________________________________

________________________________________________________________

---

Q38 Have you been to other in-person conferences since March 2020?

◯ Yes (1)

◯ No (2)

◯ Prefer not to answer (3)

---

*Display This Question:*

*If Have you been to other in-person conferences since March 2020? = Yes*

Q39 Were your behaviors and perceived risk similar or different? Please elaborate.

________________________________________________________________

________________________________________________________________

________________________________________________________________

________________________________________________________________

________________________________________________________________

Q40 Which individual provisions did you feel were helpful in reducing the risk of contracting an airborne infection like COVID-19?

- ▢ N95 masks (1)
- ▢ Access COVID-19 surveillance information about Bethlehem (2)
- ▢ Access to free rapid tests (3)
- ▢ Access to basic public health guidance information (4)
- ▢ Free PCR tests (6)
- ▢ Other (5) ________________________________________________

Q41 Which structural provisions did you feel were helpful in reducing the risk of contracting an airborne infection like COVID-19?

- ▢ Encouraging and making freely accessible N95 masks to everyone (1)
- ▢ Encouraging and making freely accessible rapid tests everyone (2)
- ▢ Reducing attendee density via hybrid option (3)
- ▢ Air ventilation and measurement (4)
- ▢ Provision of COVID relief funds/hybrid presentation option for those who test positive before or during the conference (5)
- ▢ Upper Air UV Lamp fixtures (7)
- ▢ Other (6) ________________________________________________
- ▢ Prefer not to answer (8)

---

Q42 Did any COVID mitigations seem unnecessary, intrusive, obtrusive, or counterproductive? If so, how?

________________________________________________________________

________________________________________________________________

________________________________________________________________

________________________________________________________________

________________________________________________________________

---

Q43 How did you perceive other people's behavior around COVID precautions during the conference? Please feel free to share any general impressions, but please do not identify anybody individually or by name.

________________________________________________________________

________________________________________________________________

________________________________________________________________

________________________________________________________________

________________________________________________________________

Q44  Is there anything you would like to add about your experience at the conference? If so please share here.

________________________________________________________________

________________________________________________________________

________________________________________________________________

________________________________________________________________

________________________________________________________________

End of Block: Post Conf Qual

Start of Block: Demographic Questions

Q24 Please type your age below in numbers (i.e., "48"). If you do not want to reply, please type "prefer not to answer"

________________________________________________________________

Q25 With which gender do you identify?

- ◯ Man  (1)
- ◯ Woman  (2)
- ◯ Non-binary / third gender  (3)
- ◯ Prefer to self-describe (Please specify)  (4) __________________________________________________
- ◯ Prefer not to answer  (5)

Q26 What is your race / ethnicity? (Please select all that apply)

- [ ] Asian / Pacific Islander (1)
- [ ] Black / African American (2)
- [ ] Hispanic / Latino (3)
- [ ] Native American / American Indian (4)
- [ ] White / Caucasian (5)
- [ ] Prefer to self-describe (Please specify) (6) ________________________________________________
- [ ] Prefer not to answer (7)

---

Q27 What is your highest level of education?

- ○ High School Diploma (1)
- ○ Bachelor's Degree (2)
- ○ Master's Degree (3)
- ○ Doctorate (4)
- ○ Other (Please specify) (5) ________________________________________________
- ○ Prefer not to answer (6)

---

Q28 What is your professional status?

- Tenure track, Assistant (1)
- Tenure track, Associate (2)
- Tenure track, Professor (3)
- Graduate Student (4)
- Non-tenure track (5)
- Prefer to self-describe (Please specify) (6) ________________________________________________
- Prefer not to answer (7)

End of Block: Demographic Questions
