## Appendix D: Sensitivity Estimates for "Can a Conference be Epidemiologically Conscious? A Pilot Study of Implementing COVID-19 Mitigation Measures at an In-Person Conference"

Mean sensitivity calculation assumed the following possible sensitivities for participants RAT results, derived from Soni et al [47]:

**Conditions** For each participant, test day *x* symptomatic/asymptomatic status *y* and time between the test for the present day and past two 48-hour periods to test sensitivity mapping was assigned based on day x. If participants did not respond to survey questions regarding symptoms for the day, they were counted as asymptomatic.

Let $x$ = day, of conference/post-conference (1-9) Let $\text{Day}_{\text{test}}[x]$ = 1 if a participant reported testing, 0 if they reported not testing. Let $\text{D}[x]_{sx}$ =1 if a participant reported $>$=1 symptoms associated with COVID-19; 0, if no symptoms were reported, or if no response.

For each day $x$, the following algorithm was applied based on whether a participant had a single test, two or three serials.

**Single test** $\text{Day}_{\text{test}}[x] = 1$; $\text{D}[x]_{sx}$ = 0 Single test asymptomatic: 38%

$\text{Day}_{\text{test}}[x] = 1$; $\text{D}[x]_{sx}$ = 1 Single test symptomatic: 89%

**Two Serial tests** $\text{Day}_{\text{test}}[x]$ & $\text{Day}_{\text{test}}[x-2]$ = 1; $\text{D}[x]_{sx}$ = 0 Two tests, 48 hours apart, asymptomatic 63%

$\text{Day}_{\text{test}}[x]$ & $\text{Day}_{\text{test}}[x-2]$ =1; $\text{D}[x]_{sx}$ = 0 Two tests, 48 hours apart, symptomatic 93%

**Three serial tests** $\text{Day}_{\text{test}}[x]$ & $\text{Day}_{\text{test}}[x-2]$ & $\text{Day}_{\text{test}}[x-4]$=1; $\text{D}[x]_{sx}$ = 0 Three tests, 48 hours apart, asymptomatic: 76%

$\text{Day}_{\text{test}}[x]$& $\text{Day}_{\text{test}}[x-2]$ & $\text{Day}_{\text{test}}[x-4]$=1; $\text{D}[x]_{sx}$ = 1 Three tests, 48 hours apart, symptomatic: 93%

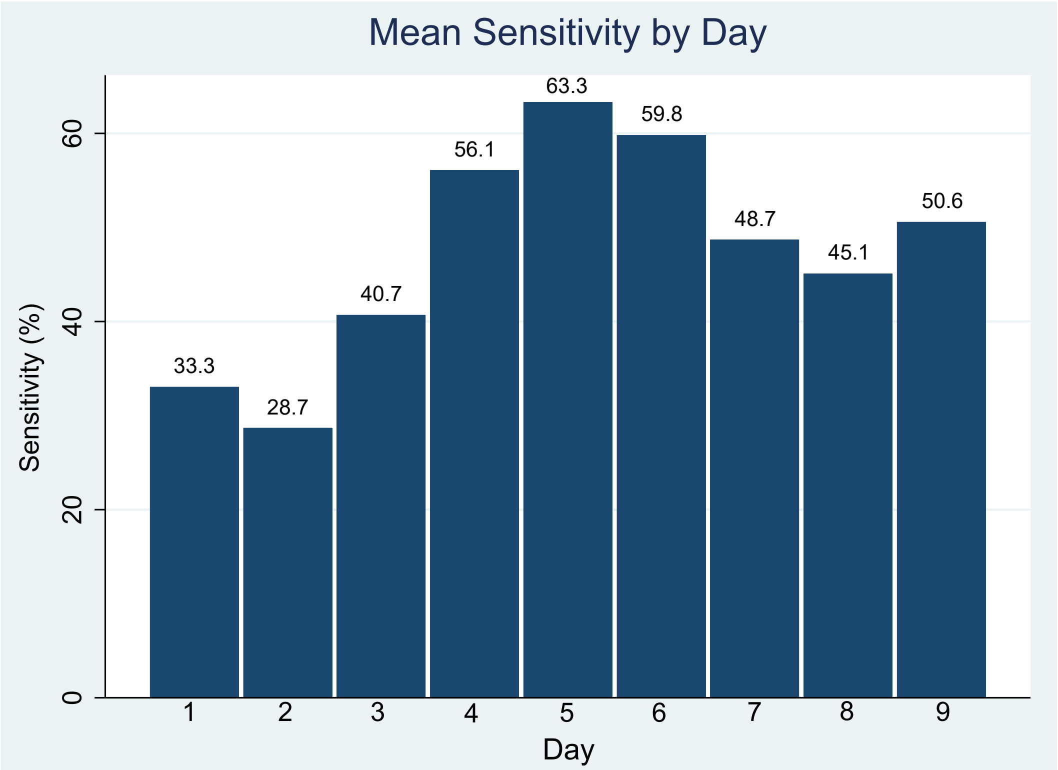


FIG. S3. The daily estimated mean sensitivity of COVID-19 rapid antigen tests for attendees. Differences in sensitivity are estimated from [47] and depend on: (i) whether the attendee has tested in the past 48 and (ii) past 96 hours, and on the symptomatic/asymptomatic status of the attendee for each test.

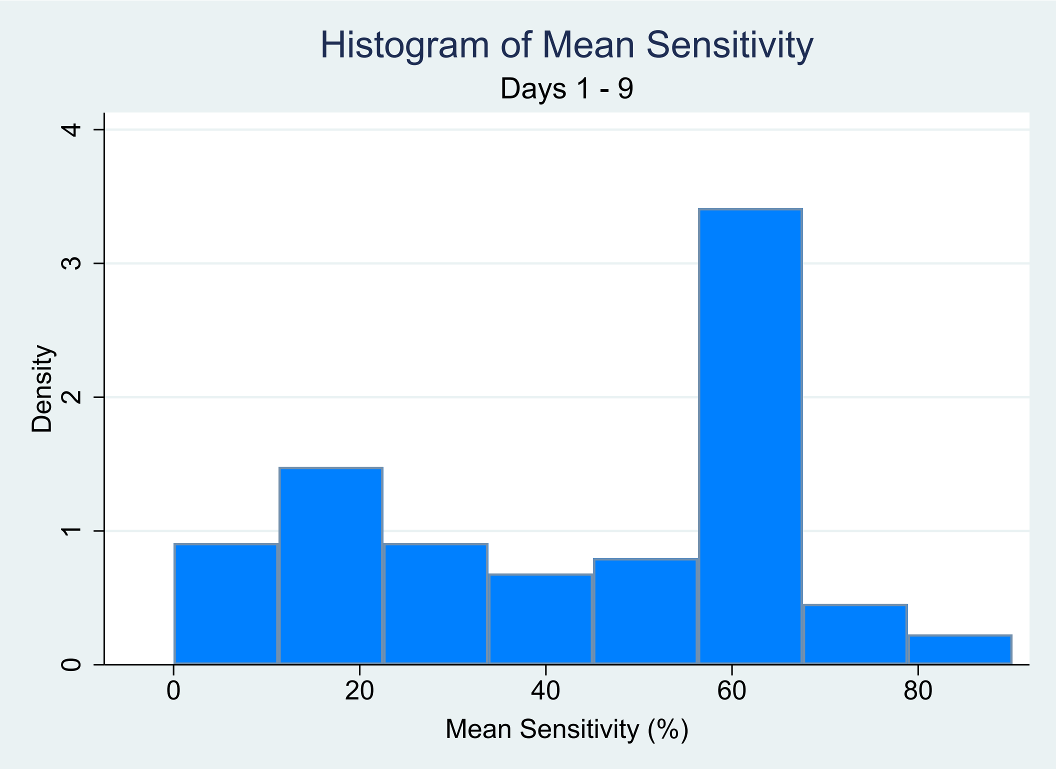


FIG. S4. Frequency distribution of mean sensitivity. Differences in sensitivity are estimated from [47] and depend on: (i) whether the attendee has tested in the past 48 and (ii) past 96 hours, and on the symptomatic/asymptomatic status of the attendee for each t
