## Appendix E: Budget for "Can a Conference be Epidemiologically Conscious? A Pilot Study of Implementing COVID-19 Mitigation Measures at an In-Person Conference"

| Item/Expense | Unit | Unit Cost | Total |
|---|---|---|---|
| 3M AURA N95s | 195 | 1.53 | 127.45 |
| FlowFlex SARS-COV-2 Antigen Rapid Test | 376 | 5.99 | 2,252.00 |
| Custom Corsi-Rosenthal Box: CO2 sensors and auto switch 10 | 10 | 49.84 | 498.84 |
| Lasko air-circulating 20-inch box fan | | | |
| Filtrete 20x20x1 Air Filter MPR 1900 MERV 13 | 20 | 22.95 | 459.00 |
| Raspberry Pi Zero W computer (reads CO monitor, calculates estimated Air exchange per hour, triggers box fan) | 10 | 15.00 | 150.00 |
| Aranet 4 CO2 Monitor | 10 | 150.00 | 1,500.00 |
| HiLetgo 2pcs 5V One-Channel Relay Module Relay Switch | 10 | 3.20 | 32.00 |
| Duct tape | 4 | 8.20 | 32.80 |
| | **Materials total** | | 5,052.90 |
| COVID Relief Aid/Registration Refund Registration cancellation refunds prior to conference due to positive test | 3 | 235 | 1,067.40 |
| Registration refund and travel relief due to positive test during conference | 1 | 362.40 | 362.40 |
| | **Materials total** | | 1,429.80 |
| | **Mitigations total** | | 6,482.70 |

TABLE S1. Itemized budget of COVID Conscious mitigations
